# A case series of SARS-CoV-2 reinfections caused by the variant of concern Gamma in Brazil

**DOI:** 10.1101/2021.11.29.21266109

**Authors:** Felipe Gomes Naveca, Valdinete Alves Nascimento, Fernanda Nascimento, Maria Ogrzewalska, Alex Pauvolid-Corrêa, Mia Ferreira Araujo, Ighor Arantes, Érika Lopes Rocha Batista, Alessandro Leonardo Álvares Magalhães, Fernando Vinhal, Tirza Peixoto Mattos, Irina Riediger, Maria do Carmo Debur, Beatriz Grinsztejn, Valdiléa G Veloso, Patricia Brasil, Rodrigo Ribeiro Rodrigues, Darcita Buerger Rovaris, Sandra Bianchini Fernandes, Cristiano Fernandes, João Hugo Abdalla Santos, Lígia Fernandes Abdalla, Rubens Costa-Filho, Marineide Silva, Victor Souza, Ágatha Araújo Costa, Matilde Mejía, Maria Júlia Brandão, Luciana Fé Gonçalves, George Allan Silva, Michele Silva de Jesus, Karina Pessoa, André de Lima Guerra Corado, Debora Camila Gomes Duarte, Ana Beatriz Machado, Ketiuce de Azevedo Zukeram, Natalia Valente, Renata Serrano Lopes, Elisa Cavalcante Pereira, Luciana Reis Appolinario, Alice Sampaio Rocha, Luis Fernando Lopez Tort, Tsuyoshi Sekizuka, Kentaro Itokawa, Masanori Hashino, Makoto Kuroda, Gabriel Luz Wallau, Edson Delatorre, Tiago Gräf, Marilda Mendonça Siqueira, Gonzalo Bello, Paola Cristina Resende

**Affiliations:** Laboratório de Ecologia de Doenças Transmissíveis na Amazônia, Instituto Leônidas e Maria Deane, Fiocruz, Manaus, Brazil; Laboratory of Respiratory Viruses and Measles, Oswaldo Cruz Institute (IOC), Oswaldo Cruz Foundation (FIOCRUZ), Rio de Janeiro, RJ, Brazil; Department of Veterinary Integrative Biosciences, Texas A&M University, College Station, Texas, United States; Laboratório de AIDS e Imunologia Molecular, Instituto Oswaldo Cruz, FIOCRUZ, Rio de Janeiro, Brazil; Secretaria de Saúde de Aparecida de Goiânia, Goiás, Brazil; Laboratório HLAGYN, Goiânia, Goiás, Brazil; Laboratório Central de Saúde Pública do Amazonas (LACEN-AM) Manaus, Amazonas, Brazil; Laboratório Central de Saúde Pública de Paraná (LACEN-PR) Curitiba, Paraná, Brazil; Instituto Nacional de Infectologia Evandro Chagas (INI), Fiocruz, Rio de Janeiro, Brazil; Laboratório Central de Saúde Pública do Espírito Santo (LACEN-ES), Vitória, Espirito Santo, Brazil; Laboratório Central de Saúde Pública do Estado de Santa Catarina (LACEN-SC), Florianópolis, Santa Catarina, Brazil; Fundação de Vigilância em Saúde do Amazonas - Dra Rosemary Costa Pinto, Manaus, Amazonas, Brazil; Hospital Adventista de Manaus, Manaus, Amazonas, Brazil; Universidade do Estado do Amazonas, Manaus, Brazil; Hospital Pró-cardíaco - Rede Américas UHG; CENUR Litoral Norte, Universidad de la República, Salto, Uruguay; Pathogen Genomics Center, National Institute of Infectious Diseases 1-23-1 Toyama, Shinjuku-ku, Tokyo 162-8640, Japan; Instituto Aggeu Magalhães, Fundação Oswaldo Cruz, Recife, Pernambuco, Brazil; Departamento de Biologia Centro de Ciências Exatas, Naturais e da Saúde, Universidade Federal do Espírito Santo, Alegre, Brazil; Instituto Gonçalo Moniz, Fundação Oswaldo Cruz, Salvador, Brazil

**Keywords:** COVID-19, SARS-CoV-2, reinfection, Variant of Concern, Gamma, P.1, Brazil

## Abstract

The rapid spread of the SARS-CoV-2 Variant of Concern (VOC) Gamma during late 2020 and early 2021 in Brazilian settings with high seroprevalence raised some concern about the potential role of reinfections in driving the epidemic. Very few cases of reinfection associated with the VOC Gamma, however, have been reported. Here we describe 25 cases of SARS-CoV-2 reinfection confirmed by real-time RT-PCR twice within months apart in Brazil. SARS-CoV-2 genomic analysis confirmed that individuals were primo-infected between March and December 2020 with distinct viral lineages, including B.1.1, B.1.1.28, B.1.1.33, B.1.195 and P.2, and then reinfected with the VOC Gamma between 3 to 12 months after primo-infection. The overall mean cycle threshold (Ct) value of the first (25.7) and second (24.5) episodes were roughly similar for the whole group and 14 individuals displayed mean Ct values < 25.0 at reinfection. Sera of 14 patients tested by plaque reduction neutralization test after reinfection displayed detectable neutralizing antibodies against Gamma and other SARS-CoV-2 variants (B.1.33, B.1.1.28 and Delta). All individuals have milder or no symptoms after reinfection and none required hospitalization. The present study demonstrates that the VOC Gamma was associated with reinfections during the second Brazilian epidemic wave in 2021 and raised concern about the potential infectiousness of reinfected subjects. Although individuals here analyzed failed to mount a long-term sterilizing immunity, they developed a high anti-Gamma neutralizing antibody response after reinfection that may provide some protection against severe disease.

## Introduction

Cases of severe acute respiratory syndrome coronavirus 2 (SARS-CoV-2) reinfection are a concerning topic for the coronavirus disease 2019 (COVID-19) response worldwide as they have implications for the long-term protective immunity induced by natural infections or by vaccines. Several cases of reinfection with distinct variants of SARS-CoV-2 have been reported^1^, but it is unclear if these reinfection cases were the consequence of a limited immunity induced by the primo-infection or reflect the reinfecting virus’s ability to evade the previous immune responses. The state of Amazonas, located in the north region of Brazil, was severely hit by a second wave of COVID-19 epidemic between December 2020 and March 2021 despite the high (>70%)^2^ or relatively high (30-35%)^3^ estimated seroprevalence at late 2020. This was associated with the emergence and spread of the Variant of Concern (VOC) Gamma which raised some concern about the potential role of reinfections in driving the second COVID-19 wave in the Amazonas^4^.

Gamma is one of the four SARS-CoV-2 VOCs currently recognized in the world^5^. All VOCs carry several mutations in the receptor-binding domain (RBD) of the spike (S) protein that reduce antibody neutralization and/or increase affinity for ACE2 receptor^6-10^, and may thus enhance the probability of reinfection. Some cases of reinfection with VOCs have been described worldwide^11-18^, but the precise frequency of those events remains unclear. A study conducted in the United Kingdom (UK) identified possible reinfections in 0.7% (95% CI 0.6-0·8%) of individuals infected during September-December 2020 and found no evidence that the frequency of reinfections was higher for the VOC Alpha than for pre-existing non-VOCs^19^. In Brazil, previous studies documented a few cases of reinfection with lineages B.1.1.28, B.1.1.33 and B.1.2^20-22^, with lineage P.2 that harbors the mutation S: E484K^23,24^ and with the VOC Gamma^17,18^. However, there is no evidence that reinfections with Gamma were more frequent than with non-VOCs.

In this study, we report a series of 25 cases of reinfections with the VOC Gamma in subjects from six different Brazilian states who had been primo-infected with different non-VOC SARS-CoV-2 lineages between 3 to 12 months earlier. The large number of SARS-CoV-2 reinfections with the VOC Gamma here described demonstrate that this viral variant is able to infect recovered individuals and suggests that this event was not a rare phenomenon in Brazil.

## Methods

### Case series and ethical aspects

In this study we include 25 cases of adults living at four different regions of Brazil, including West-Central (n=13), South (n=7), North (n=3) and Southeast (n=2) that presented two episodes of COVID-19 with at least 90 days apart. The first and second episodes occurred between March and December 2020 and between December 2020 and June 2021, respectively. All patients had nasopharyngeal and oropharyngeal swabs (NPS) collected in viral transport media (VTM) and tested by SARS-CoV-2 real time RT-PCR in their respective State Health Departments as part of the official network of the Brazilian Ministry of Health for the diagnostic and surveillance of SARS-CoV-2. Samples of patients who were SARS-COV-2 positive by real time RT-PCR twice with at least 90 days apart, according to the Technical Note 52/2020-CGPNI/DEIDT/SVS/MS^25^, were sent to the National Reference Laboratory for reinfection investigation and confirmation. This study was approved by the Ethics Committee of the Amazonas State University (CAAE: 25430719.6.0000.5016) and by the Ethics Committee of the FIOCRUZ (CAAE: 68118417.6.0000.5248).

### SARS-CoV-2 real time RT-PCR confirmation and genomic sequencing

After the detection of two SARS-CoV-2 positive samples by real time RT-PCR from the same patient the samples were accessed and sent to one of the sequencing hubs of the COVID-19 Fiocruz Genomic Surveillance Network (LVRS, Fiocruz, Rio de Janeiro; ILMD, Fiocruz Amazonas; or HLAGyn, Goiás) to have the total nucleic acid extracted from the VTM specimens by Maxwell® RSC Viral Total Nucleic Acid Purification Kit (Promega, Madison, WI) or QIAmp RNA viral mini kit (Qiagen). Then, immediately submitted to a real-time RT-PCR designed to amplify gene N of SARS-CoV-2^26^ or EDx kit Biomanguinhos protocol to amplify the gene E^27^. Using the nucleic acid extracts, we generated the whole genome of SARS-CoV-2 by the in-house amplicon sequencing protocols described ^28,29^ but with some improvements in the primer scheme (**Supplementary file**) or by the Illumina COVIDSeq test kit (Illumina) with some adaptations^30^. Libraries were produced with Nextera XT and sequenced with MiSeq Reagent Micro Kit v2 (300-cycles). The FASTQ reads were obtained following the Illumina pipeline on BaseSpace, imported into Geneious v10.2.6, trimmed (BBDuk 37.25) or into CLC Genomic Workbench (Qiagen), and mapped (BBMap 37.25) against the reference sequence EPI_ISL_402124 available in EpiCoV database from GISAID (https://www.gisaid.org/). Consensus sequences with a mean read depth of 1,341x were generated after excluding duplicate reads.

### Genomic analyses

PANGO lineages were assigned to all sequences by the Pangolin algorithm ^31^, and later confirmed using maximum likelihood (ML) phylogenetic analyses. SARS-CoV-2 complete genome sequences from all cases were aligned with high quality (<1% of N) SARS-CoV-2 whole-genomes (>29 kb) of representative lineages retrieved from EpiCoV database at GISAID. The final dataset of each variant to which primoinfection and reinfection sequences were previously assigned were obtained by clusterization of their GISAID complete datasets using CD-HIT v.4.8.1^31^.The resulting dataset was aligned by MAFFT v7.467^32^ and subjected to a maximum likelihood (ML) phylogenetic analysis with IQ-TREE v2.1.2^33^ under the best nucleotide substitution model selected by the ModelFinder application^34^. The branch support was assessed by the approximate likelihood-ratio test based on the Shimodaira–Hasegawa-like procedure (SH-aLRT) with 1,000 replicates ^35^.

### Serological analyses

A total of 14 serum samples collected from 12 to 70 days after the second episode of COVID-19 were tested for SARS-CoV-2-specific neutralizing antibodies (NAb) by plaque reduction neutralization test (PRNT_90_). We also include serum samples from two control groups: 1) samples collected 3 to 21 days after hospitalization from 30 individuals primo-infected with the Gamma variant and 2) samples collected from 10 individuals with hybrid immunity acquired by fully vaccination after infection with a non-VOC variant (n = 5) or by infection with the Gamma variant after vaccination. For PRNT_90_, an aliquot of serum sample inactivated at 56°C for 30 minutes was tested in VERO CCL-81 cells in duplicate at serial two-fold dilutions to determine 90% endpoint titers against four infectious SARS-CoV-2 lineages, including the LVRS reference strains B.1.1.33 (EPI_ISL_1181439), B.1.1.28 (EPI_ISL_2645638), Gamma (EPI_ISL_1402431) and Delta (EPI_ISL_2645417). Serum samples were considered seropositive when a serum dilution of at least 1:10 reduced no less than 90% of the formation of SARS-CoV-2 viral plaques^36^.

### Statistical analyses

The non-parametric Wilcoxon matched-pairs signed rank test was used to compare multiple samples per subject (real time RT-PCR cycle threshold [Ct] of samples from the first and second episodes and level of NAb against different SARS-CoV-2 variants in the plasma taken after reinfection) and the Mann-Whitney test was used to compare samples from different groups of individuals. Only Ct values from samples analyzed with the same real time RT-PCR diagnostic assay were compared. Specimens in which NAb could not be detected (PRNT_90_ <10) were assigned an arbitrary value of five in order to include NAb as a continuous variable. The threshold for statistical significance was set to *P* < 0.05. Graphics and statistical analyses were performed using GraphPad v9.02 (Prism Software, United States).

## Results

We analyzed 25 individuals that presented two episodes of COVID-19 within an interval of 3 to 12 months period (**Table 1, Figure S1**). They were predominantly female (64%), unvaccinated (92%) and with an age that ranged from 17 to 73 years old. Most cases had no reported comorbidities (80%) and presented mild clinical symptoms (92%) including fever, myalgia, cough, sore throat, nausea, anosmia, ageusia, and back pain in the first episode of COVID-19. Two individuals required hospitalization at primo-infection. Patients had milder clinical presentation (84%) or were asymptomatic (16%) during the second infection. All samples tested were positive to SARS-CoV-2 by real time RT-PCR with a Ct value ranging from 18.0 to 34.3. The overall mean Ct value of the first (25.7) and second (24.5) episodes were not significantly different (*P* > 0.05) for the whole group (**Fig 1A**). Of note, 14 individuals displayed mean Ct values < 25.0 during the second episode of COVID-19 and nine individuals displayed much lower Ct values in the second than in the first episode (Ct_first_ - Ct_second_ > 3.0). The overall mean between onset symptoms and collection date were 4 days in the first infection and 3.5 in the reinfection (**Table 1**).

**Table 1.**
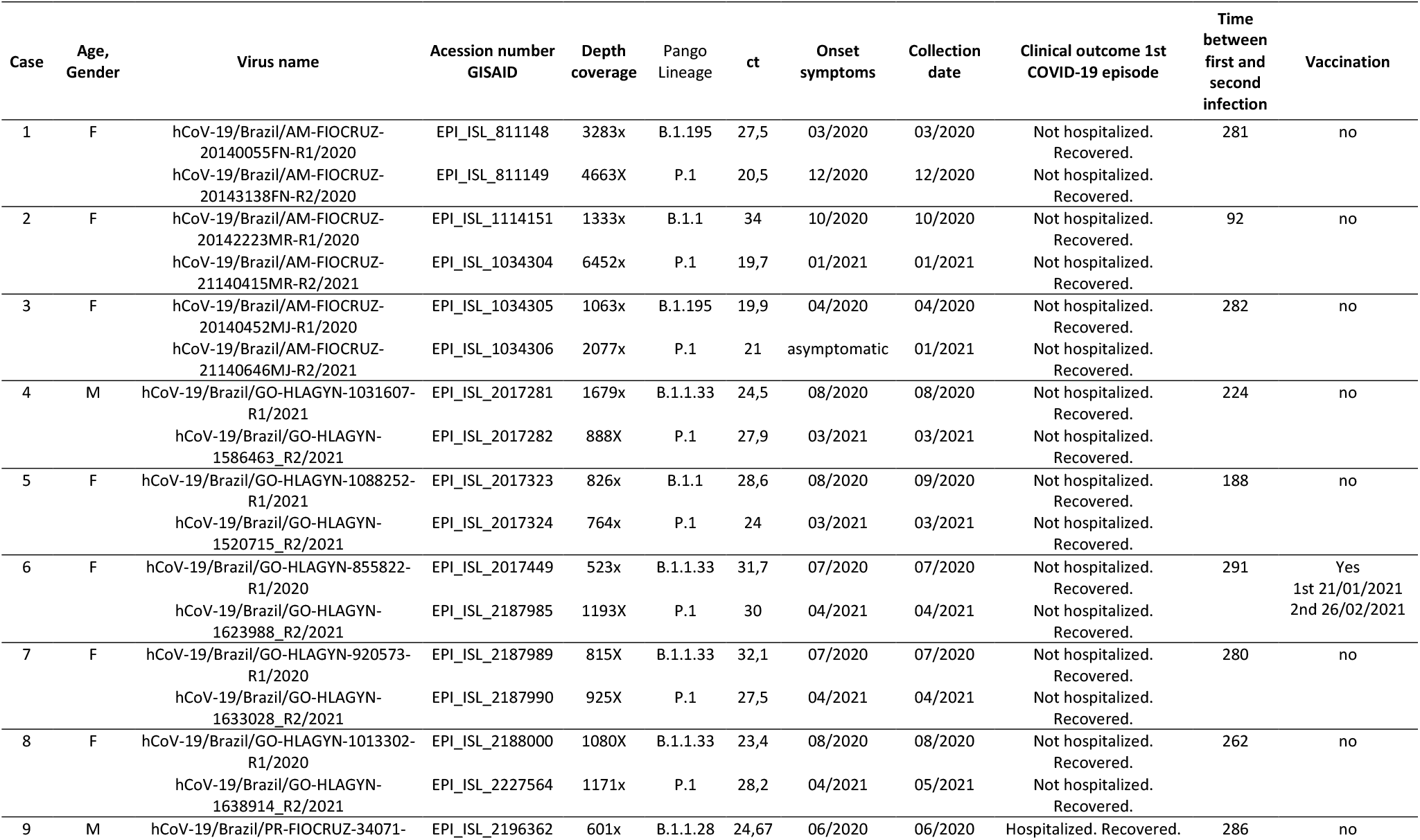
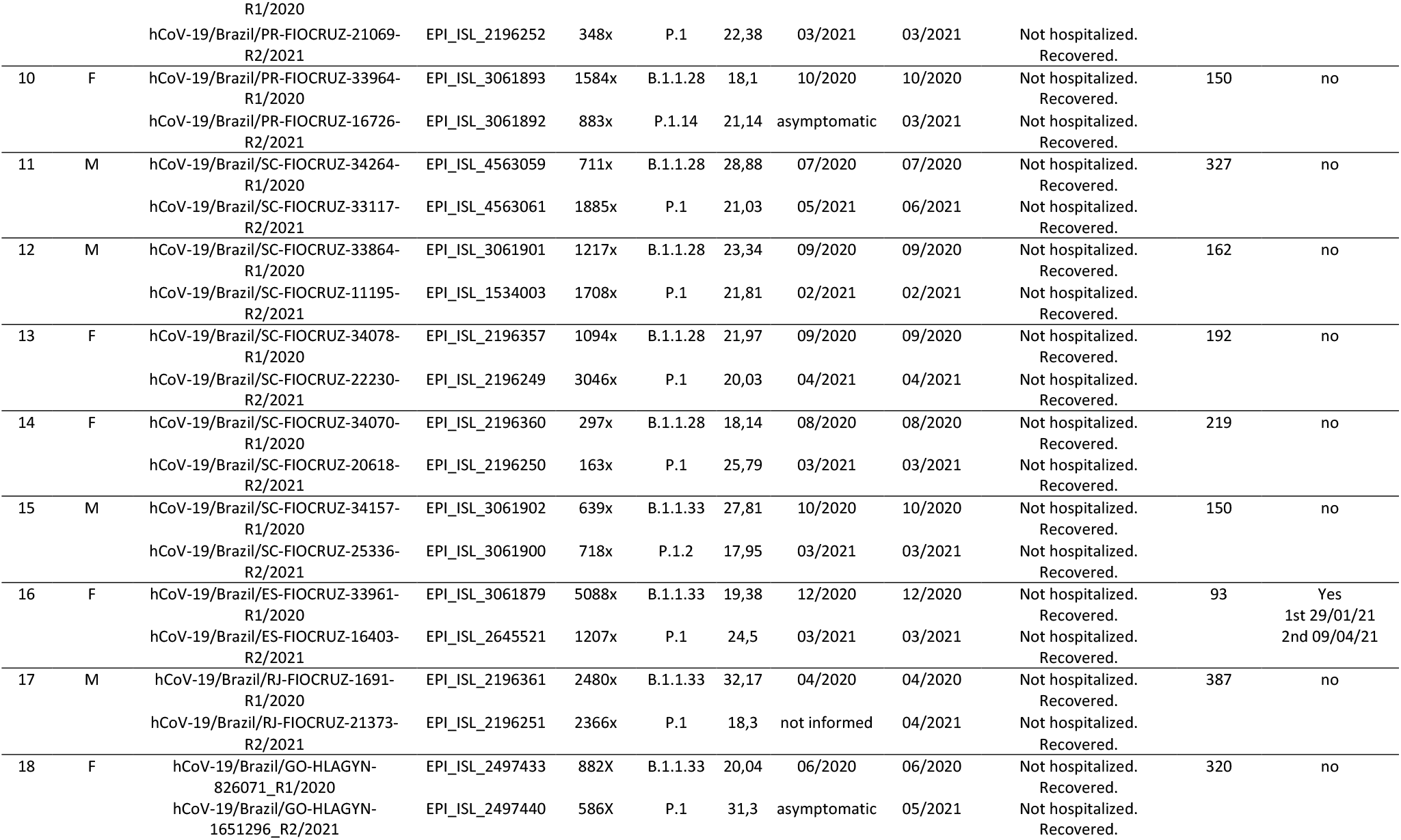
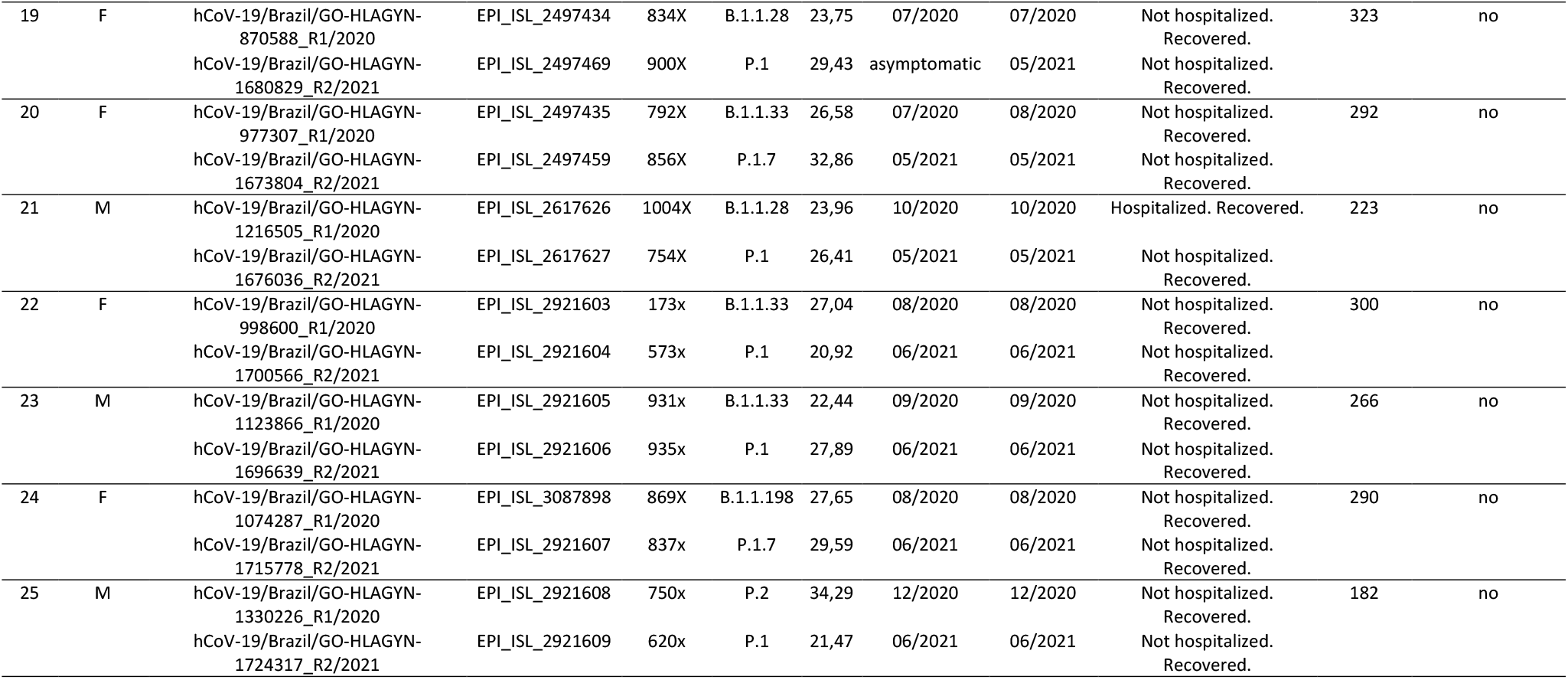
Clinical and epidemiological details of Gamma reinfection cases.

**Figure 1.**
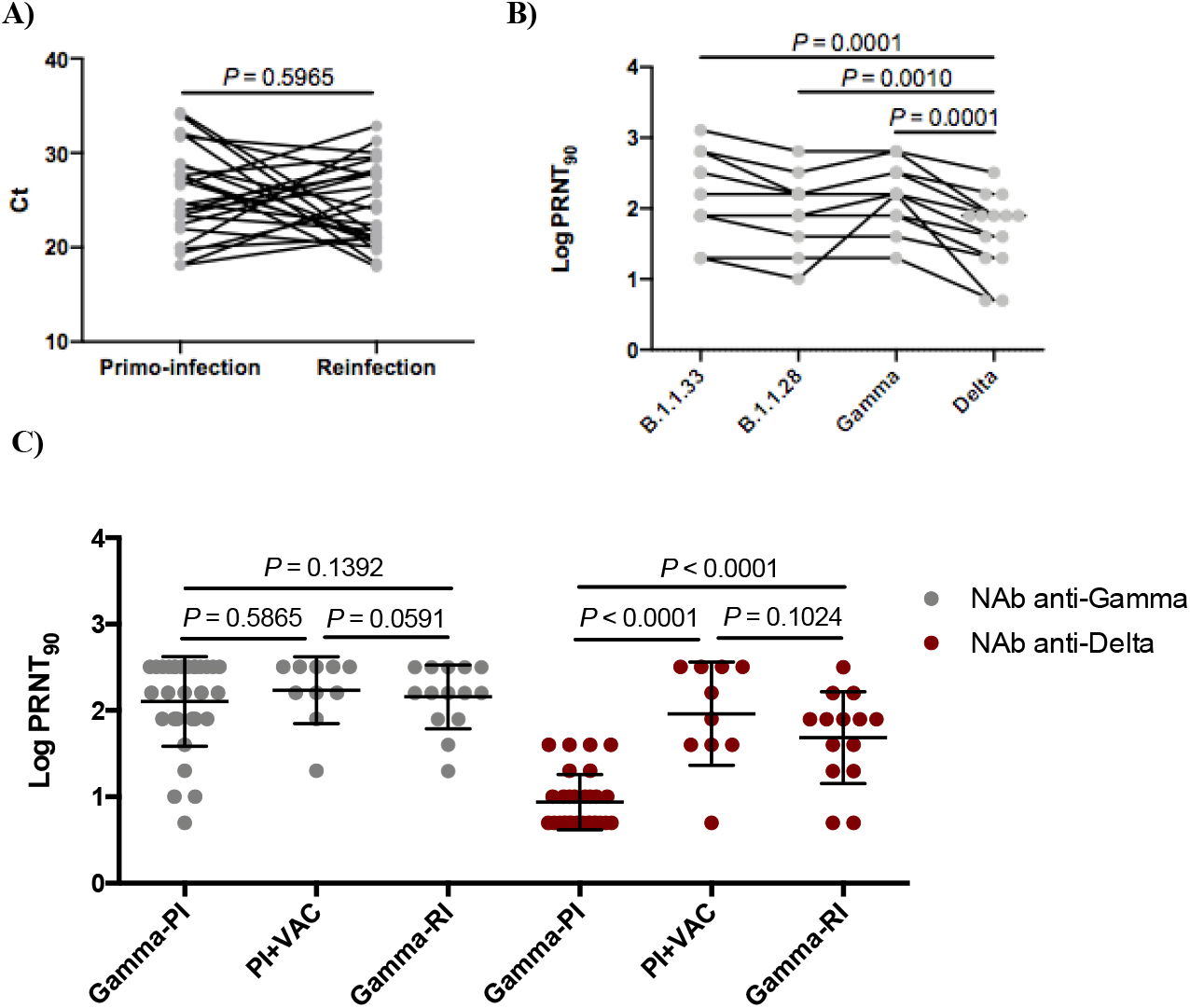
Viral load in NSP samples and neutralization capacity of convalescent serum of reinfected individuals. **A)** Real time RT-PCR Ct value distribution of NSP SARS-CoV-2 positive samples taken from 25 individuals at primo-infection and reinfection. Each line represents a single individual. **B)** Individual trajectories of neutralization titers (PRNT_90_) against different SARS-CoV-2 lineages in convalescent plasma from 14 individuals collected 10-75 days following SARS-CoV-2 reinfection. Each line represents a single individual. **C)** Comparison of neutralization titers (PRNT_90_) against Gamma and Delta variants in convalescent plasma from three groups that included: 1) hospitalized individuals primo-infected with Gamma (*n* = 30; Gamma-PI), 2) individuals with hybrid immunity acquired by vaccination after infection or by infection after vaccination (*n* = 10; PI+VAC), and 3) individuals reinfected with Gamma (*n* = 14; Gamma-RI). Horizontal bars represent sample medians and interquartile range. Two-tailed *P* values calculated with the Wilcoxon matched-pairs signed rank test (A and B) or the Mann-Whitney test (C) are shown.

Whole SARS-CoV-2 genomes were recovered from all 50 samples analyzed. Most of them were high quality sequences (<1% of N) with a few exceptions (<15% of N) that were recovered from samples with low viral load (Ct > 33), limit Ct value for sequencing procedure. All SARS-CoV-2 genomes recovered contained enough SNP markers to confidently assign the corresponding SARS-CoV-2 lineage with high support (1.0). The PANGO lineage system indicated the presence of five different SARS-CoV-2 lineages in the first COVID-19 episodes (B.1.1, B.1.1.28, B.1.1.33, B.1.195, B.1.1.198 and P.2) and the unique presence of the VOC Gamma (lineage P.1) and Gamma plus (lineages P.1.2, P.1.7, P.1.12 and P.1.14) in all second episodes (**Table 1**). The ML phylogenetic analysis confirmed the initial lineage assignment on the majority of cases (**Fig S2 A and B**). A single divergence (EPI_ISL_1114151, Patient 2) was detected in a genome with a significant number of unidentified positions (14%) that could potentially lead to ambiguity in its classification. This allowed us to conclude that all suspected cases correspond to reinfections with the VOC Gamma. Analysis of the Spike (S) gene of Gamma viruses detected at reinfection reveals the presence of the canonical lineage signatures plus additional mutations L5F and T76I in one P.1 sequence, D178G in one P.1.12 sequence, A522V in one P.1 sequence and P681H in two P.1.7 sequences (**Table S1, Figure S2 C**).

Serum samples of 14 patients collected 10-75 days after the second SARS-CoV-2 positive real time RT-PCR were tested for plaque reduction neutralization against B.1.1.28, B.1.1.33, Gamma and Delta variants. Selected individuals were primo-infected with lineages B.1.1 (*n* = 3), B.1.1.28 (*n* = 4), B.1.1.33 (*n* = 5), B.1.195 (*n* = 1), and P.2 (*n* = 1). Most patients (86%) have detectable NAb against all viral lineages tested, although the neutralization geometric mean titer against Delta (PRNT_90_ = 49) was significantly lower (*P* < 0.05) than for B.1.1.28 (PRNT_90_ = 103), B.1.1.33 (PRNT_90_ = 160) and Gamma (PRNT_90_ = 160) (**Fig 1B and Table 1**). The levels of Nab in reinfected subjects were next compared with a group of 30 hospitalized individuals that were primo-infected with variant Gamma and 10 individuals with hybrid immunity that were either primo-infected with a non-VOC variant and then fully vaccinated (*n* = 5) or infected with the variant Gamma after vaccination (*n* = 5, breakthrough cases). The three groups displayed similar levels of NAb against Gamma (*P* > 0.05), but levels against Delta in reinfected and in individuals with hybrid immunity were significantly higher (*P* < 0.0003) than in hospitalized individuals primo-infected with Gamma (**Fig 1C)**.

## Discussion

This study describes 25 individuals from Brazil that were primo-infected with SARS-CoV-2 lineages B.1.195 (two cases), B.1.1 (two cases), B.1.1.28 (eight cases), B.1.1.33 (11 cases), B.1.1.198 (one case) and P.2 (one case) between March and December 2020 and reinfected with the VOC Gamma 3-12 months later (between December 2020 and June 2021). Lineages B.1.1.28 and B.1.1.33 were the most prevalent Brazilian variants between March and October 2020, lineage B.1.195 was locally prevalent in the Amazonas state from March to June 2020, lineage B.1.1 circulates at low prevalence in Brazil during 2020, lineage P.2 was highly prevalent in most Brazilian states between November 2020 and February 2021 and the VOC Gamma was frequently detected in the Amazonas state since December 2020 and became the most prevalent viral variant across all Brazilian regions from February to July 2021 (http://www.genomahcov.fiocruz.br/dashboard/). Thus, the viruses detected at primo-infection and reinfection in our study reflect the most prevalent contemporaneous SARS-CoV-2 variants circulating in Brazil at the time period.

These cases of reinfection as well as those previously reported for the variants P.2/Gamma in Brazil^17,23,24^ are consistent with the described capability of viruses carrying the substitution S:E484K to escape from anti-SARS-CoV-2 antibodies^7-9^ and to cause SARS-CoV-2 breakthrough infections in vaccinated persons^37-39^. These observations are also consistent with previous studies that showed that variant Gamma is more resistant to neutralization by sera from convalescent/vaccinated patients than ancestral Wuhan related strains^40-45^. It is possible that the absence of anti-SARS-CoV-2 antibodies against the S: K484 variant in subjects infected during 2020 may have facilitated reinfections with variants Gamma and P.2. The detection of one case of reinfection with Gamma following primo-infection with lineage P.2; however, supports the hypothesis that reinfection might have also occurred in spite of pre-existing anti-S: K484 antibodies. It is also possible that the large number of detected reinfection cases by the VOC Gamma here detected resulted from its high infectivity and transmissibility phenotype ^46,47^.

We may also speculate that mild severity of the first COVID-19 episode in patients here described induced a transient protective immunity, but anti-SARS-CoV-2 NAb substantially decayed by the time of reinfection^48-51^. One previous study that provided estimates of the typical time period to reinfection for several coronaviruses, and predicted that reinfection with SARS-CoV-2 under endemic conditions would likely occur between 3 and 63 months after peak antibody response, with a median of 16 months^52^. Although we do not measure the anti-SARS-CoV-2 NAb levels before reinfections, all individuals in our study were reinfected between 3 and 12 months after the primo-infections, a time-frame thus consistent with the hypothesis of waning humoral immunity. According to this model, reinfection will become increasingly common as the epidemic progresses, and this may also partially explain the much larger number of reinfections with variant Gamma here reported when compared with previous locally prevalent variants in Brazil.

There is limited evidence on the risk of SARS-CoV-2 transmission from reinfected individuals to susceptible contacts. Several studies demonstrated that Ct values that inversely correlate with the log viral load also negatively correlate with cultivable virus^53,54^ and the transmission risk^55,56^. It was reported that up to 70% of patients remained positive in culture at a Ct ≤ 25^53,54^ and that 85% of case-contact pairs with plausible onward transmission had a case Ct < 25 ^55,56^, suggesting that a Ct ≤ 25 could be used as a good surrogate of infectivity. Notably, our analyses revealed that samples taken at first and second infections displayed roughly similar mean Ct values (25.7 *vs* 24.5) and that 56% (14/25) of individuals displayed a mean Ct value < 25.0 at reinfection. These findings support that, individuals here analyzed display comparable viral loads in the upper respiratory tract at primo-infection and reinfection and that viral load at reinfection may have been sufficient for some patients to transmit the virus to others. Thus, reinfections with the VOC Gamma might have contributed to the onward transmission of SARS-CoV-2 in Brazil, although the precise frequency of those events remains unknown.

In the present study, the serum samples of 14 patients were collected between 10-75 days after the reinfection and all of them displayed detectable NAb against Gamma and also against the other three SARS-CoV-2 lineages tested (B.1.1.28, B.1.1.33 and Delta). It is also interesting to note that most patients here analyzed were young (<50 years old), have no known underlying conditions and all of them have a benign clinical outcome during the second infection. These findings suggest that reinfections did not occur because of genetic immunological deficits in the individuals here analyzed and that partial immunity induced by early SARS-CoV-2 variants may have provided some protection against severe illness during reinfection with the variant Gamma^57^. Furthermore, the Gamma reinfected subjects have similar levels of NAb against Gamma and Delta variants than a group of individuals with hybrid immunity (fully vaccination after infection or breakthrough cases) and higher levels against Delta than a group of subjects primo-infected with Gamma, supporting that reinfection may have boosted neutralizing titers against different viral variants as seen after vaccination of previously infected persons^58-60^.

Longitudinal analyses revealed that anti-SARS-CoV-2 IgG levels and *in vitro* NAb titers induced by natural infection correlate with protection against reinfection ^61-64^ but the precise level of NAb necessary for protection from infection and/or reinfection remains unclear^65^. Of interest, sera from patients here collected 10-75 days after reinfection were less potent to neutralize the variant Delta, relative to the variant Gamma and other non-VOCs tested. This observation coincides with a recent study that revealed that sera from convalescents individuals infected with VOCs Beta and Gamma showed a mark reduction of ∼11-fold in neutralization of Delta compared with the corresponding infecting VOCs ^66^ and with another study that points that SARS-CoV-2 variants can elicit polyclonal antibodies with different immunodominance hierarchies^67^. Considering the recent and ongoing dissemination of variant Delta in Brazil and the progress of vaccination roll-out, it will be important to determine whether individuals primo-infected with Gamma that were vaccinated or not will be at risk of reinfection with Delta.

Our study has several limitations. First, although Ct values could be used as a surrogate of infectivity, we have no contact-tracing information to demonstrate that reinfected subjects were able to transmit the virus to susceptible individuals. Second, our study was limited by the absence of longitudinal data of anti-SARS-CoV-2 NAb, particularly between the first and the second COVID-19 episodes. Thus, we could not confirm if primo-infections effectively induce a transient neutralization response, we could not measure the level of NAb immediately before reinfections and no definitive conclusions could be drawn about the precise impact of reinfections on NAb titers.

In summary, these findings confirm that natural infection by SARS-CoV-2 do not necessarily prevent subsequent infections and further demonstrate that reinfection with the VOC Gamma in people who had a first symptomatic infection between 3 to 12 months earlier was not a rare phenomenon in Brazil. Although, reinfections cases were observed in individuals without comorbidities and with mild disease, no significant decrease in viral load, represented by Ct values, during reinfection was observed when compared with primo-infection. Further studies are necessary to determine whether reinfection with Gamma and other VOCs is a widespread phenomenon and to understand to what extent reinfection will contribute to the onward endemic transmission of SARS-CoV-2. Further work is also needed to understand how primary infections interfere with the clinical outcome of secondary infections and the degree of long-term sterilizing immunity acquired after reinfections.

### Role of the funding source

The sponsors of the study had no role in study design, data collection, data analysis, data interpretation, or writing of the report and in the decision to submit the paper for publication. All authors confirm that they had full access to all the data in the study and accept responsibility to submit for publication.

## Data Availability

The consensus SARS-CoV-2 sequences generated in this work are available online at EpiCoV database in GISAID https://www.gisaid.org under the accession numbers: EPI_ISL_811148, EPI_ISL_811149, EPI_ISL_1034304 to 1034306, EPI_ISL_1114151, EPI_ISL_1534003, EPI_ISL_2017281 to 2017324, EPI_ISL_2017449, EPI_ISL_2187985, EPI_ISL_2187989, EPI_ISL_2187990, EPI_ISL_2188000, EPI_ISL_2196249 to 2196252, EPI_ISL_2196357, EPI_ISL_2196360 to 2196362, EPI_ISL_2227564, EPI_ISL_2497433 to 2497435, EPI_ISL_2497440, EPI_ISL_2497459, EPI_ISL_2497469, EPI_ISL_2617626, EPI_ISL_2617627, EPI_ISL_2645521, EPI_ISL_2921603 to 2921609, EPI_ISL_3061879, EPI_ISL_3061892, EPI_ISL_3061893, EPI_ISL_3061900 to 3061902, EPI_ISL_3087898, EPI_ISL_4563059 and EPI_ISL_4563061.

## Contributors

FGN, AAM and MMS obtaining financial support. FGN, MMO, APC, IIA, GGB and PCR contributed to data analysis and writing of the manuscript. AAM, ERB, TPM, IIR, MCD, BBG, VGV, PPB, RRR, DBR, SBF, CCF, JAS, LFA and RCF contributed to patient and public health surveillance data. AAC, AGC, ASR, DBR, DGD, ECP, FFV, FGN, FN, GAS, IIR, KAZ, KKP, LFG, LRA, MCD, MJB, MMM, MMO, MMS, MSJ, PCR, RRR, RSL, SBF, TPM, VAN and VVS contributed to diagnostics and genomic sequencing. ABM, APC, LLT, MFA and NNV contributed to viral isolation and neutralization analyses. EED, GGB, GLW, IIA and TTG performed the phylogenetic analysis. TS, KI, MH, and MK contributed with editing of the manuscript and sharing previously unpublished data. All authors read and accept the participation in this manuscript.

## Declaration of interests

All authors declare no competing interests.

## Funding

Funding support from Department of Science and Technology (DECIT), Ministry of Health (MoH), CGLab/MoH (General Laboratories Coordination of Brazilian Ministry of Health), CVSLR/FIOCRUZ (Coordination of Health Surveillance and Reference Laboratories of Oswaldo Cruz Foundation), CNPq COVID-19 MCTI 402457/2020-0 and 403276/2020-9; INOVA Fiocruz VPPCB-005-FIO-20-2 and VPPCB-007-FIO-18-2-30; FAPERJ: E26/210.196/2020; FAPEAM (PCTI-EmergeSaude/AM call 005/2020 and Rede Genômica de Vigilância em Saúde - REGESAM). This study was also supported by a Grant-in Aid from the Japan Agency for Medical Research and Development (AMED) under Grant number JP20fk0108103.

## Acknowledgments

We would like to thank all Genomic Coronavirus Fiocruz Network members (http://www.genomahcov.fiocruz.br/), the Multi-user Research Facility of Biosafety Level 3 Platform of the Oswaldo Cruz Institute (IOC), Fiocruz, and the CGLab/MoH and Secretary of Surveillance and Health of the Brazilian MoH (SVS-MS). Additionally, we gratefully acknowledge the Authors from the originating laboratories responsible for obtaining the specimens and the submitting laboratories where genetic sequence data were generated and shared via the GISAID, on which this research is based (**Supplementary Table 1**).

## Supplementary Figures

**Figure S1:**
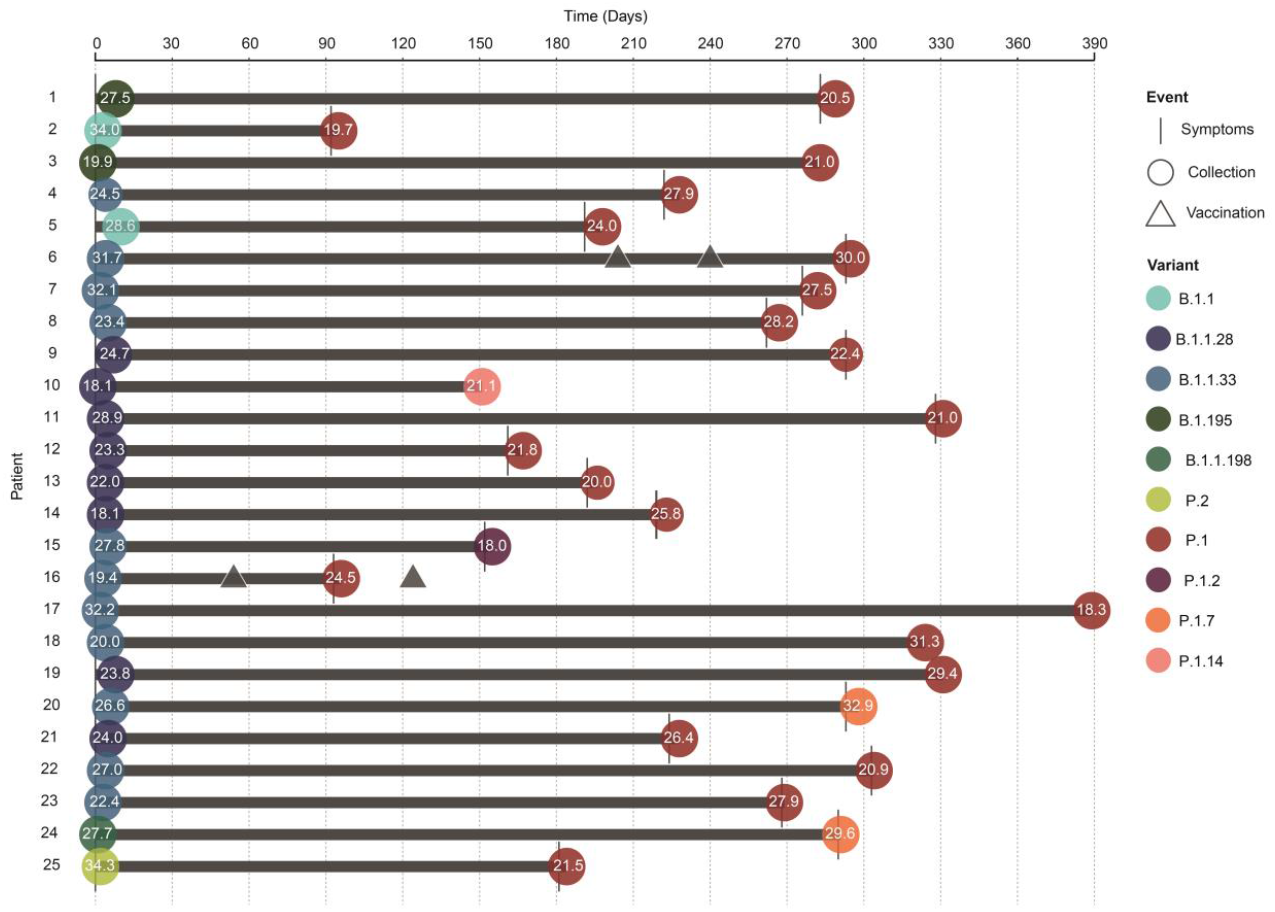
Epidemiological History of Gamma Reinfection Cases. The graph illustrates the epidemiological history of Gamma reinfection cases (*n* = 25), detailing its key events. When available, the onset of symptoms and sample collection for primo-infection and reinfection cases are represented in a timeline drawn for each individual. Sample collection events are accompanied by their observed real time PCR cycle threshold (Ct). Additionally, vaccination events are indicated. All time intervals are calculated from the onset of symptoms of the individual’s primo-infection.

**Figure S2.**
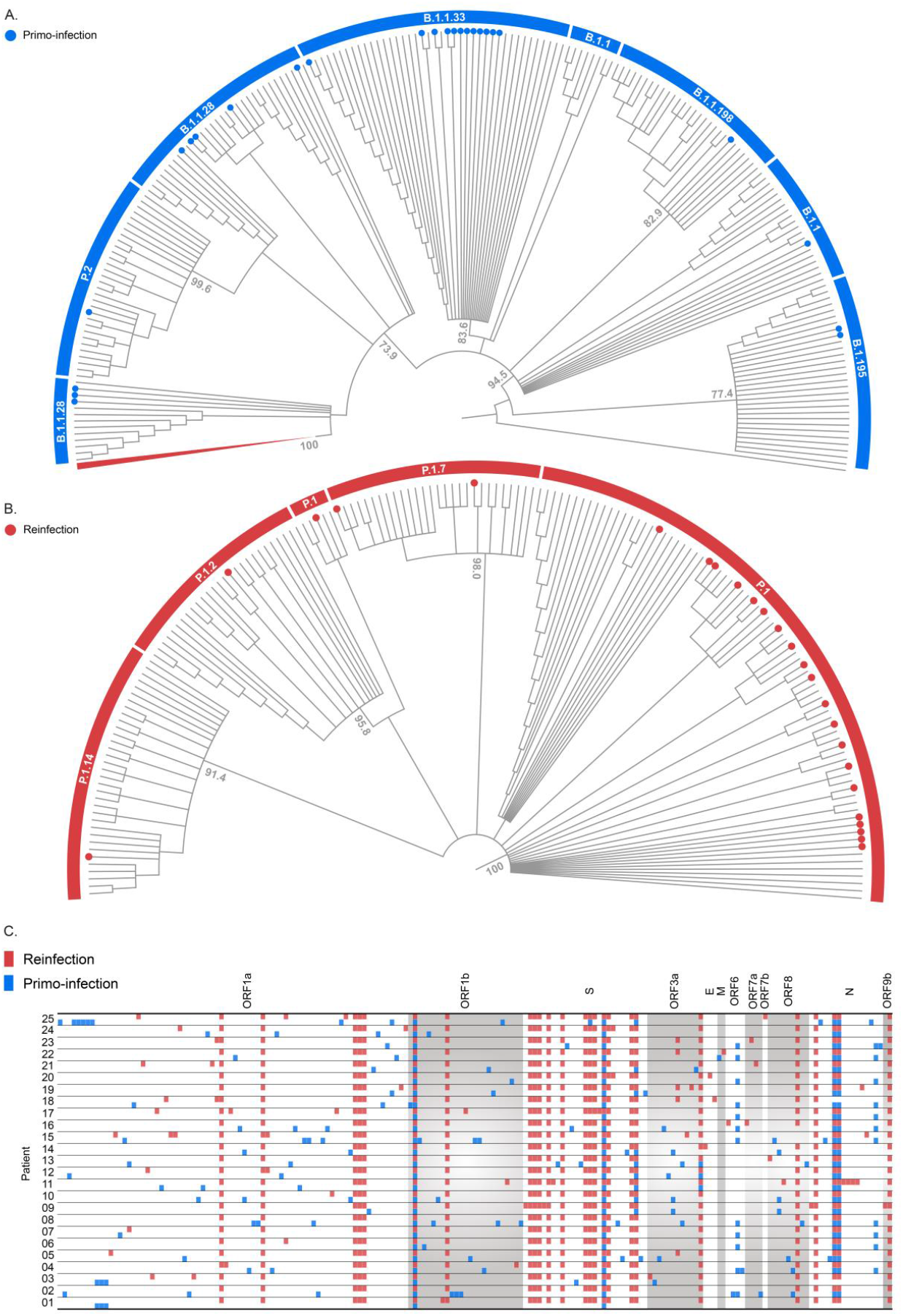
Phylogenetic and Molecular Characterization of Gamma Reinfection Cases. The figure details the divergence observed between the SARS-CoV-2 genomes associated to each individual’s primo-infection and reinfection cases. A. Cladogram-transformed Maximum likelihood (ML) tree (*n* = 375) of complete genomes (29,605 *nts*) from a selected group of variants identified in the outside blue archway. Primo-infection sequences (n = 25) are indicated by blue circles. The branch leading to the Gamma (P1 + P1*) variant is collapsed and colored in red. The tree was rooted using Wuhan reference sequence (EPI_ISL_402124). B. Cladogram-transformed ML tree of the Gamma variant subtree (*n* = 172) collapsed in A. Gamma variants are identified in the outside red archway. Reinfection cases (*n* = 25) are colored in red. Statistical support (SH-aLRT) of key nodes in A and B are indicated in the trees. C. Mapping of observed mutations in primo-infection (blue squares) and reinfection (reds squares) genomes for all patients (*n* = 25).

## Notes

### Competing Interest Statement

The authors have declared no competing interest.

### Funding Statement

Funding support from Department of Science and Technology (DECIT), Ministry of Health, CGLab/MoH (General Laboratories Coordination of Brazilian Ministry of Health), CVSLR/FIOCRUZ (Coordination of Health Surveillance and Reference Laboratories of Oswaldo Cruz Foundation), CNPq COVID-19 MCTI 402457/2020-0 and 403276/2020-9; INOVA Fiocruz VPPCB-005-FIO-20-2 and VPPCB-007-FIO-18-2-30; FAPERJ: E26/210.196/2020; FAPEAM (PCTI-EmergeSaude/AM call 005/2020 and Rede Genomica de Vigilancia em Saude - REGESAM). This study was also supported by a Grant-in Aid from the Japan Agency for Medical Research and Development (AMED) under Grant number JP20fk0108103.

### Author Declarations

This study was approved by the Ethics Committee of the Amazonas State University (CAAE: 25430719.6.0000.5016) and by the Ethics Committee of the FIOCRUZ (CAAE: 68118417.6.0000.5248).

