## Supplemental Table 1 for "A case series of SARS-CoV-2 reinfections caused by the variant of concern Gamma in Brazil"

We gratefully acknowledge the following Authors from the Originating laboratories responsible for obtaining the specimens, as well as the Submitting laboratories where the genome data were generated and shared via GISAID, on which this research is based.

All Submitters of data may be contacted directly via www.gisaid.org

Authors are sorted alphabetically.

| Accession ID | Originating Laboratory | Submitting Laboratory | Authors |
|---|---|---|---|
| EPI_ISL_1082291 | "Stefan S. Nicolau"Institute of Virology | "Stefan S. Nicolau"Institute of Virology | Adriana Plesa; Alina Nastasie; Ana Iulia Neagu; Anca Botezatu; Camelia Sultana; Carmen Cristina Diaconu; Coralia Bleotu; Cristina Mambet; Denisa Dragu; Gabriela Anton; Ioana Pitica; Iulia Virginia Iancu; Laura Grecu; Laura Necula; Lilia Matei; Marinela Bostan; Mihaela Economescu; Mirela Mihaila; Saviana Nedeianu; Simona Ruta |
| EPI_ISL_2493746 | AFIP | Instituto Butantan | Antonio Jorge Martins; Claudia Renata dos Santos Barros; David Schlesinger; Debora Botequio Moretti; Dimas Tadeu Covas; Elaine Cristina Marqueze; Elaine Vieira Santos; Evandra Strazza Rodrigues; Heidge Fukumasu; Jayme Augusto de Souza-Neto; José Salvatore Leister Patané; Luiz Alcantara; Luiz Lehmann Coutinho; Maria Carolina Elias; Maurício Lacerda Nogueira; Rafael dos Santos Bezerra; Raul Machado Neto; Rejane Maria Tommasini Grotto; Ricardo Haddad; Sandra Coccuzzo Sampaio Vessoni; Simone Kashima; Svetoslav Nanev Slavov; Vincent Louis Viala |
| EPI_ISL_2494061 | AFIP SUDESTE | Instituto Butantan | Antonio Jorge Martins; Claudia Renata dos Santos Barros; David Schlesinger; Debora Botequio Moretti; Dimas Tadeu Covas; Elaine Cristina Marqueze; Elaine Vieira Santos; Evandra Strazza Rodrigues; Heidge Fukumasu; Jayme Augusto de Souza-Neto; José Salvatore Leister Patané; Luiz Alcantara; Luiz Lehmann Coutinho; Maria Carolina Elias; Maurício Lacerda Nogueira; Rafael dos Santos Bezerra; Raul Machado Neto; Rejane Maria Tommasini Grotto; Ricardo Haddad; Sandra Coccuzzo Sampaio Vessoni; Simone Kashima; Svetoslav Nanev Slavov; Vincent Louis Viala |
| EPI_ISL_2494083 | AFIP SUL | Instituto Butantan | Antonio Jorge Martins; Claudia Renata dos Santos Barros; David Schlesinger; Debora Botequio Moretti; Dimas Tadeu Covas; Elaine Cristina Marqueze; Elaine Vieira Santos; Evandra Strazza Rodrigues; Heidge Fukumasu; Jayme Augusto de Souza-Neto; José Salvatore Leister Patané; Luiz Alcantara; Luiz Lehmann Coutinho; Maria Carolina Elias; Maurício Lacerda Nogueira; Rafael dos Santos Bezerra; Raul Machado Neto; Rejane Maria Tommasini Grotto; Ricardo Haddad; Sandra Coccuzzo Sampaio Vessoni; Simone Kashima; Svetoslav Nanev Slavov; Vincent Louis Viala |
| EPI_ISL_500681 | Area of Virology, Serology and Virology Division (SAViD), New South Wales Health Pathology Randwick | Area of Virology, Serology and Virology Division (SAViD), New South Wales Health Pathology Randwick | Rawlinson, W. |
| EPI_ISL_2493764 | BIOFAST | Instituto Butantan | Antonio Jorge Martins; Claudia Renata dos Santos Barros; David Schlesinger; Debora Botequio Moretti; Dimas Tadeu Covas; Elaine Cristina Marqueze; Elaine Vieira Santos; Evandra Strazza Rodrigues; Heidge Fukumasu; Jayme Augusto de Souza-Neto; José Salvatore Leister Patané; Luiz Alcantara; Luiz Lehmann Coutinho; Maria Carolina Elias; Maurício Lacerda Nogueira; Rafael dos Santos Bezerra; Raul Machado Neto; Rejane Maria Tommasini Grotto; Ricardo Haddad; Sandra Coccuzzo Sampaio Vessoni; Simone Kashima; Svetoslav Nanev Slavov; Vincent Louis Viala |
| EPI_ISL_2494230 | BIOFAST LESTE | Instituto Butantan | Antonio Jorge Martins; Claudia Renata dos Santos Barros; David Schlesinger; Debora Botequio Moretti; Dimas Tadeu Covas; Elaine Cristina Marqueze; Elaine Vieira Santos; Evandra Strazza Rodrigues; Heidge Fukumasu; Jayme Augusto de Souza-Neto; José Salvatore Leister Patané; Luiz Alcantara; Luiz Lehmann Coutinho; Maria Carolina Elias; Maurício Lacerda Nogueira; Rafael dos Santos Bezerra; Raul Machado Neto; Rejane Maria Tommasini Grotto; Ricardo Haddad; Sandra Coccuzzo Sampaio Vessoni; Simone Kashima; Svetoslav Nanev Slavov; Vincent Louis Viala |
| EPI_ISL_2345874 | CENTRO DE APOIO EPIDEMIOLOGICO | Instituto Butantan / Mendelics | Antonio Jorge Martins; Claudia Renata dos Santos Barros; David Schlesinger; Debora Botequio Moretti; Dimas Tadeu Covas; Elaine Cristina Marqueze; Elaine Vieira Santos; Evandra Strazza Rodrigues; Heidge Fukumasu; Jayme Augusto de Souza-Neto; José Salvatore Leister Patané; Luiz Alcantara; Luiz Lehmann Coutinho; Maria Carolina Elias; Maurício Lacerda Nogueira; Rafael dos Santos Bezerra; Raul Machado Neto; Rejane Maria Tommasini Grotto; Ricardo Haddad; Sandra Coccuzzo Sampaio Vessoni; Simone Kashima; Svetoslav Nanev Slavov; Vincent Louis Viala |
| EPI_ISL_1795299 | CENTRO DE SAUDE III AFFONSO LUZZI SANTA CRUZ DAS PALMEIRAS | Instituto Butantan / ESALQ-Piracicaba | Antonio Jorge Martins; Bianca Cechetto Carlos. Mendelics: Bibiana Santos; Claudia Renata dos Santos Barros; David Schlesinger. Hemocentro Ribeirão Preto: Simone Kashima; Debora Botequio Moretti. Centro de Genômica Funcional da ESALQ: Luiz Lehmann Coutinho; Dimas Tadeu Covas; Elaine Cristina Marqueze; Elaine Vieira dos Santos; Elisangela Chicaroni Mattos; Erika Freitas; Evandra Strazza Rodrigues; Felipe Allan da Silva da Costa; Flavia Aburjaile; Guilherme Targino Valente; Heidge Fukumasu. USP-Botucatu: Rejane Maria Tommasini Grotto; Instituto Butantan: Alexander Roberto Precioso; Jayme A. Souza-Neto; Jessika Cristina Chagas Lesbon; José Salvatore Leister Patané; João Paulo Kitajima; Luiz Carlos Junior de Alcantara; Maria Carolina Elias; Marta Giovanetti; Patricia Akemi Assato; Rafael dos Santos Bezerra; Raquel de Lello Rocha Campos Cassano. NGS Soluções Genômicas: Pilar Drummond Sampaio Corrêa Mariani. FZEA-USP Pirassununga: Mirele Daiana Poleti; Raul Machado Neto; Ricardo Augusto Brassaloti; Ricardo Haddad; Rodrigo Tocantins Calado.; Sandra Coccuzzo Sampaio; Svetoslav Nanev Slavov; Vagner Fonseca; Vincent Louis Viala |
| EPI_ISL_2344700 | CENTRO MEDICO DR NELSON SALOME DE CONCHAL | Instituto Butantan / FZEA-USP-Pirassununga | Antonio Jorge Martins; Claudia Renata dos Santos Barros; David Schlesinger; Debora Botequio Moretti; Dimas Tadeu Covas; Elaine Cristina Marqueze; Elaine Vieira Santos; Evandra Strazza Rodrigues; Heidge Fukumasu; Jayme Augusto de Souza-Neto; José Salvatore Leister Patané; Luiz Alcantara; Luiz Lehmann Coutinho; Maria Carolina Elias; Maurício Lacerda Nogueira; Rafael dos Santos Bezerra; Raul Machado Neto; Rejane Maria Tommasini Grotto; Ricardo Haddad; Sandra Coccuzzo Sampaio Vessoni; Simone Kashima; Svetoslav Nanev Slavov; Vincent Louis Viala |
| EPI_ISL_3758170, EPI_ISL_3758183, EPI_ISL_4554787 | CHECK-UP - Medicina e Diagnóstico - Matriz | HLAGYN - Laboratorio de Imunologia de Transplantes de Goias | Elaize Maria Gomes de Paula; Fernando Antonio Vinhal dos Santos; Frederico Rodrigues Vinhal; Gladstone Rodrigues da Cunha Filho; Kamila Oliveira Reis De Freitas; Lucas Carlos Gomes Pereira; Nubia Silva Araújo; Sabrina Sara Moreira Duarte |
| EPI_ISL_815260 | Centogene | Centogene | Krishna Kumar Kandaswamy; Peter Bauer; Vivi Hue-Trang Lieu |
| EPI_ISL_978503, EPI_ISL_978510, EPI_ISL_978517, EPI_ISL_978518, EPI_ISL_978526, EPI_ISL_1068321 | Central Public Health Laboratory - LACEN - Bahia, Salvador, Brazil | Central Public Health Laboratory - LACEN - Bahia, Salvador, Brazil | Arabela Leal; Breno Dominguez; Felicidade Pereira; Jaqueline Gomes; Luciana Oliveira; Luiz Alcantara; Marcela Gómez; Marta Giovanetti; Patrícia Cajado; Stephane Tosta; Vagner Fonseca; Vanessa Nardy |
| EPI_ISL_1468430 | Centro de Saude II Matao | Instituto Adolfo Lutz, Interdiciplinary Procedures Center, Strategic Laboratory | Caio Vinicius Dias Lopes; Claudia Regina Gonçalves; Claudio Tavares Sacchi; Erica Valessa Ramos Gomes; Karoline Rodrigues Campos |
| EPI_ISL_497864 | Department of Microbiology, The University of Hong Kong | Department of Microbiology, The University of Hong Kong | Kelvin K.W. To; Kwok-Yung Yuen |
| EPI_ISL_855558, EPI_ISL_855559 | Department of Virology, Principal Military Hospital of Instruction of Tunis | Bundeswehr Institute of Microbiology | Habiba Naija; Kilian Stoecker; Malena Bestehorn-Willmann; Markus H. Antwerpen; Mathias C. Walter; Roman Wölfel & Mohamed Ben Moussa; Simone Eckstein; Susann Handrick |
| EPI_ISL_2344544 | ESALQ | Instituto Butantan / ESALQ-Piracicaba | Antonio Jorge Martins; Claudia Renata dos Santos Barros; David Schlesinger; Debora Botequio Moretti; Dimas Tadeu Covas; Elaine Cristina Marqueze; Elaine Vieira Santos; Evandra Strazza Rodrigues; Heidge Fukumasu; Jayme Augusto de Souza-Neto; José Salvatore Leister Patané; Luiz Alcantara; Luiz Lehmann Coutinho; Maria Carolina Elias; Maurício Lacerda Nogueira; Rafael dos Santos Bezerra; Raul Machado Neto; Rejane Maria Tommasini Grotto; Ricardo Haddad; Sandra Coccuzzo Sampaio Vessoni; Simone Kashima; Svetoslav Nanev Slavov; Vincent Louis Viala |
| EPI_ISL_1469572, EPI_ISL_1469661 | FUNDACAO DE SAUDE PUBLICA DE NOVO HAMBURGO FSNH | Epiclin | Ana Paula Muterle; Carolina Comerlato; Eliana Márcia Da Ros Wendland; Fernando Hayashi Sant'Anna; Janira Prichula; Juliana Comerlato |
| EPI_ISL_1182551 | Fundação Ezequiel Dias (FUNED) | Coordenação Geral de Laboratórios de Saúde Pública (CGLAB/DAEVS/SVS/MS) | Vagner Fonseca; et al. |
| EPI_ISL_1278015 | GA Department of Public Health | GA Department of Public Health | Aliyah Fields; Cynthia Dixey; Jonathan Edwards; Stacy Reeves; Taylor Smith; Tonia Parrott |
| EPI_ISL_861911 | Genomika Einstein | LATE - Laboratório de Técnicas Especiais - Hospital Israelita Albert Einstein | Ana Paula Moreira Salles; Deyvid Amgarten; Fernanda de Mello Malta; João Bosco Oliveira Filho; João Renato Rebello Pinho; Pedro Henrique Sebe Rodrigues; Raquel Riyuzo |
| EPI_ISL_2017250, EPI_ISL_2017256, EPI_ISL_2017263, EPI_ISL_2017272, EPI_ISL_2017281, EPI_ISL_2017282, EPI_ISL_2017319, EPI_ISL_2017323, EPI_ISL_2017324, EPI_ISL_2017329, EPI_ISL_2017334, EPI_ISL_2017337, EPI_ISL_2017449, EPI_ISL_2187697, EPI_ISL_2187716, EPI_ISL_2187720, EPI_ISL_2187737, EPI_ISL_2187745, EPI_ISL_2187753, EPI_ISL_2187766, EPI_ISL_2187796, EPI_ISL_2187814, EPI_ISL_2187832, EPI_ISL_2187886, EPI_ISL_2187920, EPI_ISL_2187982, EPI_ISL_2187985, EPI_ISL_2187989, EPI_ISL_2187990, EPI_ISL_2188000, EPI_ISL_2188009, EPI_ISL_2188012, EPI_ISL_2227564, EPI_ISL_2227565, EPI_ISL_2348612, EPI_ISL_2497433, EPI_ISL_2497434, EPI_ISL_2497435, EPI_ISL_2497438, EPI_ISL_2497440, EPI_ISL_2497457, EPI_ISL_2497458, EPI_ISL_2497459, EPI_ISL_2497469, EPI_ISL_2617611, EPI_ISL_2617626, EPI_ISL_2617627, EPI_ISL_2680908, EPI_ISL_2921603, EPI_ISL_2921604, EPI_ISL_2921605, EPI_ISL_2921606, EPI_ISL_2921607, EPI_ISL_2921608, EPI_ISL_2921609, EPI_ISL_3087898, EPI_ISL_3254371, EPI_ISL_3254638, EPI_ISL_3254648, EPI_ISL_3254682, EPI_ISL_3259355, EPI_ISL_3259369, EPI_ISL_3274744, EPI_ISL_3386150, EPI_ISL_3997179, EPI_ISL_4061656, EPI_ISL_4061660 | | | |
| see above | HLAGYN - Laboratorio de Imunologia de Transplantes de Goias | HLAGYN - Laboratorio de Imunologia de Transplantes de Goias | Alessandro Leonardo Alvares Magalhaes; Daniel Ferreira de Sousa; Danielle de Paiva Rezende; Erika Lopes Rocha Batista; Fernando Antonio Vinhal dos Santos; Frederico Rodrigues Vinhal; Kamila Oliveira Reis De Freitas.; Lucas Carlos Gomes Pereira; Paola Cristina Resende Silva; Raphael Bessa Parmigiane; Sabrina Sara Moreira Duarte |
| EPI_ISL_2345023 | HOSP MUN JOSANIAS CASTANHA BRAGA | Instituto Butantan | Antonio Jorge Martins; Claudia Renata dos Santos Barros; David Schlesinger; Debora Botequio Moretti; Dimas Tadeu Covas; Elaine Cristina Marqueze; Elaine Vieira Santos; Evandra Strazza Rodrigues; Heidge Fukumasu; Jayme Augusto de Souza-Neto; José Salvatore Leister Patané; Luiz Alcantara; Luiz Lehmann Coutinho; Maria Carolina Elias; Maurício Lacerda Nogueira; Rafael dos Santos Bezerra; Raul Machado Neto; Rejane Maria Tommasini Grotto; Ricardo Haddad; Sandra Coccuzzo Sampaio Vessoni; Simone Kashima; Svetoslav Nanev Slavov; Vincent Louis Viala |
| EPI_ISL_2758676 | HUEM/IBMP | IPEC Guarapuava | NAPI-Genômica (Novos Arranjo de Pesquisa e Inovação em Genômica): Ademar Dantas da Cunha Júnior Adriano Ferrasa Adriano Mondini Aldo Przybysz Alessandra Lourenço Cecchini Armani Alex Sandro Jorge Alexandra Ivo de Medeiros Alexandre Maller Aline Cristina Batista Rodrigues Johann Ana Lucia Ferreira Ana Marisa Fusco Almeida Anderson Joel Martino Andrade André Luís Laforga Vanzela Andrea Duarte Doetzer Andrea Name Colado Simao Andressa Pereira de Souza Anelisa Ramão Angelica Beate Winter Boldt Anna Hermínia Castro Gomes de Amorim Anna Silvia Penteado Setti da Rocha Antonio Camilo da Silva Filho Antonio Stabelini Neto Arthur Hirata Bertachi Barbara Mendes Paz Chao Betty Cristiane Kuhn Bruno Ambrozio Galindo Bruno Ribeiro Cruz Camilla Reginatto De Pierri Carla Fredrichsen Moya Araujo Carla Fredrichsen Moya Araujo Carlos Alberto Oliveira de Biagi Junior Carlos Augusto Nassar Carlos Eduardo Buss Carlos Gilberto Carlotti Junior Carlos Henrique Schneider Carolina Panis Carolina Weigert Galvão Caroline de Jesus Coelho Donha Caroline Guisantes de Salvo Toni Caryna Eurich Mazur Catiuscie Cabreira da Silva Tortorella Celso F. D. Doliveira Cesar Luiz Boguszewski Christiane Pienna Soares Chung Man Chin Claudia Moro Cleverson Busso Cristiane Cominetti Daiane Priscila Simão-Silva Dalila Luciola Zanette Daniel de Paula Daniel de Paula Daniel Rech Daniela Fiori Gradia Daniela Pretti da Cunha Tirapelli Daniela Viganó Zanoti Jeronymo Daniele Ukan Danielle Malheiros Ferreira Danielle Venturini Deborah Catharine de Assis Leite Deivid Calebe de Souza Dennis Armando Bertolini Edenir Inez Pamero Edna Maria Vissoci Reiche Edson Roberto Arpini Miguel Eduardo José de Almeida Araújo Eliana Carolina Vespero Eliandro Reis Tavares Elza Kimura Grimshaw Emanuel Maltempi de Souza Emanuele Cristina Gustani Buss Émerson Carraro Emiliana Cristina Melo ENILze Maria de Souza Fonseca Ribeiro Enilze Maria de Souza Fonseca Ribeiro Erika Izumi Erika Seki Kioshima Cotica Evani Marques Pereira Fabio Negretti Fábio Rodrigues Ferreira Seiva Felipe Dunin dos Santos Felipe Tuon Fernanda Andreia Rosa Fernanda Cestaro Prado Cortez Fernanda Ivanski Fernanda Maris Peria Flavia Regina Oliveira de Barros Franciele Aní Caovilla Follador Franciele Mara Lucca Zanardo Bohm Francinete Ramos Campos Fulviana Silva Nishiyama GABRIEL RIBEIRO CORDEIRO Gabriela Datsch Bennemann Gisele Santos de Oliveira Glaucio Valdameri Glauco Akelinghton Freire Vitiello Glauco Vieira Miranda Glaura Scantamburlo ALves Fernandes Guilherme Ferreira Silveira Gustavo Bianchini Porfírio Gustavo Lenci Marques Hélito Volpato Hildebrando Masshiro Nagai Huei Diana Lee Ilce Mara de Syllos Cólus Iris Rabinovich Israel Gomy Jackson Kawakami Jacques Duílio Brancher Jaime Luís Lopes Rocha Jaqueline Carvalho de Oliveira Jean Henrique da Silva Rodrigues Jean Leandro dos Santos Jeane Eliete Laguila Visentainer João Paulo Bianchi Ximenez Joaquim Manoel da Silva Jociani Ascari Joel Donazzolo Jorge Luis Maria Ruiz Jose Knopfholz José Luis da Conceição Silva José Sebastião dos Santos Joseane Carla Schabarum Juliana Cheleski Wiggers Juliana Mara Serpeloni Juliana Morini Küpper Cardoso Perseguini Karen Brajão de Oliveira Karin Braun Prado Karine Aparecida de Lima Katiany Rizzieri Caleffi Ferracioli Katiuscia de Oliveira Francisco Gabriel Kelvinson Fernandes Viana Larissa Beatriz Cossalter Larissa Danielle Bahls Pinto Laurival Antonio Vilas Boas Léia Carolina Lucio Líbero Mezzadri Neto Ligia Carla Faccin Galhardi Lirane Elize Defante Ferreto Luciana Furlaneto Maia Luciana Oliveira de Fariña Luciana Reis Azevedo Alanis Luciane Regina Cavalli Lucy Megumi Yamauchi Lioni Luis Paulo Gomes Mascarenhas Luis Paulo Gomes Mascarenhas Luís Paulo Mascarenhas Lupe Furtado Alle Lyvia Regina Biagi Silva Bertachi Mara Antonia Ramos Costa Mara L. Cordeiro Marcela Maria Birolim Marcelo Ricardo Vicari Marcia Edilaine Lopes Consolaro Marcia Holsbach Beltrame Marcia Regina Eches Perugini Marcos Abdo Arbex Marcos Pileggi MARCOS TADEU GRZELCZAK Marcus Peikriszwili Tartaruga Maria Angelica Ehara Watanabe Maria Antonia Ramos Costa Maria Claudia Gross Maria José Soares Mendes Giannini Maria Leandra Terencio Maria Lúcia Bonfleur Maria Luiza Guimarães de Oliveira Maria Luiza Petzl-Erler Mariana Abe Vicente Cavagnari Marina Kimiko Kadowaki Marise Fonseca dos Santos Marla Karine Amarante Maurício Turkiewicz Mauro Antonio Alves Castro Michel Rodrigo Zambrano Passarini Michele Potrich Michelle Orane Schemberger Milena Massumi Kozonoe Mônica Degraf Cavallin Monica Tereza Suldofski |

| | | | |
|---|---|---|---|
| | | | Mucio Luiz de Assis Cirino Nadia Graciele Krohn Najeh Maissar Khalil Nédia de Castilhos Ghisi Neide Tomimura Costa Neiva Leite Neyva Maria Lopes Romeiro Patrícia Amâncio da Rosa Patricia Dayane Carvalho Schaker Patricia Oehlmeyer Nassar Patrícia Savio de Araújo-Souza Patricia Silva Lucio Paulo Henrique Couto Souza Paulo Roberto Donadio Percy Nohama Quirino Alves de Lima Neto Rafael Deminice Rafael dos Santos Bezerra Raquel Alves dos Santos Renan Manozzo Galante Renata Ernlund Freitas de Macedo Rita de Cássia Garcia Simão Roberta Losi Guembarovski Roberto H. Herai Roberto Rosati Rodrigo Ferreira Rodrigo Rodrigues Matiello Rogério Neri Shinsato Rogério Pincela Mateus Rosane Aparecida Ribeiro Rosilene Fressatti Cardoso Rosilene Fressatti Cardoso Sandra Mara Guse Scós Venske Selene Elifio Esposito Sérgio Ossamu Ioshii Silvana Giuliatti Silvia Mara de Souza Halick Silvio Henrique Maia de Almeida Simone Neumann Wendt Spencer Luiz Marques Payão Stefan Wolanski Negrão Stephane Janaina de Moura Escobar Sueli Fumie Yamada Ogatta SUELI PERCIO QUINAIA Taciane Finatto Tatiana Mayumi Veiga Iriyoda Tayza Katelline Danilau Ostroski Tony Alexander Hild Valeria Valente Vanessa Nascimento Kozak Vanessa Santos Sotomaior Victor Breno Pedrosa Victoria Zeghbi Cochenski Borba Vivian Rotuno Moure Valdameri Wander Rogerio Pavanelli Weber Cláudio Francisco Nunes da Silva Willian Augusto de Melo Yohandra Reyes Torres |
| EPI_ISL_861636 | Hospital Geral de Sao Mateus São Paulo | Instituto Adolfo Lutz, Interdiciplinary Procedures Center, Strategic Laboratory | Claudia Regina Gonçalves; Claudio Tavares Sacchi; Erica Valessa Ramos Gomes; Karoline Rodrigues Campos |
| EPI_ISL_414014 | Hospital Israelita Albert Einstein | Instituto Adolfo Lutz, Interdiciplinary Procedures Center, Strategic Laboratory | Carlos Henrique Camargo; Claudia Regina Gonçalves; Claudio Tavares Sacchi; Ester Cerdeira Sabino; Katia Correia dos Santos; Maria do Carmo Sampaito Tavares Timenetsky; Terezinha Maria de Paiva |
| EPI_ISL_2614570 | Hospital Sagrada Família- Unidade Maua | Instituto Adolfo Lutz, Interdiciplinary Procedures Center, Strategic Laboratory | Caio Vinicius Dias Lopes; Claudia Regina Gonçalves; Claudio Tavares Sacchi; Erica Valessa Ramos Gomes; Karoline Rodrigues Campos; Leonardo Jose Tadeu de Araujo |
| EPI_ISL_861639 | Hospital Sao Paulo de Ensino da Unifesp | Instituto Adolfo Lutz, Interdiciplinary Procedures Center, Strategic Laboratory | Claudia Regina Gonçalves; Claudio Tavares Sacchi; Erica Valessa Ramos Gomes; Karoline Rodrigues Campos |
| EPI_ISL_414017 | Hospital São Joaquim Beneficencia Portuguesa | Instituto Adolfo Lutz, Interdiciplinary Procedures Center, Strategic Laboratory | Carlos Henrique Camargo; Claudia Regina Gonçalves; Claudio Tavares Sacchi; Daniela Bernardes Borges da Silva; Ester Cerdeira Sabino; Fabiana Cristina Pereira dos Santos; Maria do Carmo Sampaito Tavares Timenetsky; Terezinha Maria de Paiva |
| EPI_ISL_861635 | Hospital e Maternidade Madre Theodora | Instituto Adolfo Lutz, Interdiciplinary Procedures Center, Strategic Laboratory | Claudia Regina Gonçalves; Claudio Tavares Sacchi; Erica Valessa Ramos Gomes; Karoline Rodrigues Campos |
| EPI_ISL_2203825 | Houston Methodist Hospital | Houston Methodist Hospital | Ilya J. Finkelstein; James J. Davis; Jessica Cambric; Jimmy Gollihar; Kristina Reppond; Layne Pruitt; Madison N. Shyer; Marcus Nguyen; Matthew Ojeda Saavedra; Paul A. Christensen; Prasanti Yerramilli; Randall J. Olsen; Robert Olson; Ryan Gadd; S. Wesley Long; Sishir Subedi; and James M. Musser |
| EPI_ISL_981387 | IAL Regional de Bauru | Instituto Adolfo Lutz, Interdiciplinary Procedures Center, Strategic Laboratory | Claudia Regina Gonçalves; Claudio Tavares Sacchi; Erica Valessa Ramos Gomes; Karoline Rodrigues Campos |
| EPI_ISL_1213196 | IMT-UFRN/RN | Bioinformatics Laboratory / LNCC | Alessandra P Lamarca; Alexandra L Gerber; Ana Paula Melo Mariano; Ana Paula de C Guimarães; Ana Tereza R Vasconcelos; Angela Maria Guimarães Santos; Bianca Mendes Maciel; Danielle Angst Secco; Eduardo Sérgio Soares Sousa; Eloiza Helena Campana; Francisco Paulo Freire Neto; George Rego Albuquerque; Kátia Castanho Scortecci; Lucymara Fassarella Agnez Lima; Luiz G P de Almeida; Luís Cristóvão Porto; Otavio J. Brustolini; Paulo Ricardo Nascimento; Ronaldo da Silva Francisco Jr; Sandra Rocha Gadelha; Selma Maria Bezerra Jeronimo; Vinícius Pietta Perez |
| EPI_ISL_2493975 | INSIDE CRSSU IPIRANGA | Instituto Butantan | Antonio Jorge Martins; Claudia Renata dos Santos Barros; David Schlesinger; Debora Botequio Moretti; Dimas Tadeu Covas; Elaine Cristina Marqueze; Elaine Vieira Santos; Evandra Strazza Rodrigues; Heidge Fukumasu; Jayme Augusto de Souza-Neto; José Salvatore Leister Patané; Luiz Alcantara; Luiz Lehmann Coutinho; Maria Carolina Elias; Maurício Lacerda Nogueira; Rafael dos Santos Bezerra; Raul Machado Neto; Rejane Maria Tommasini Grotto; Ricardo Haddad; Sandra Coccuzzo Sampaio Vessoni; Simone Kashima; Svetoslav Nanev Slavov; Vincent Louis Viala |
| EPI_ISL_2493717 | INSIDE DIAGNÓSTICOS | Instituto Butantan | Antonio Jorge Martins; Claudia Renata dos Santos Barros; David Schlesinger; Debora Botequio Moretti; Dimas Tadeu Covas; Elaine Cristina Marqueze; Elaine Vieira Santos; Evandra Strazza Rodrigues; Heidge Fukumasu; Jayme Augusto de Souza-Neto; José Salvatore Leister Patané; Luiz Alcantara; Luiz Lehmann Coutinho; Maria Carolina Elias; Maurício Lacerda Nogueira; Rafael dos Santos Bezerra; Raul Machado Neto; Rejane Maria Tommasini Grotto; Ricardo Haddad; Sandra Coccuzzo Sampaio Vessoni; Simone Kashima; Svetoslav Nanev Slavov; Vincent Louis Viala |
| EPI_ISL_2445063, EPI_ISL_2445079, EPI_ISL_2445082 | INSIDE DIAGNÓSTICOS SUDESTE VILA PRUDENTE/SAPOPEMBA | Instituto Butantan | Antonio Jorge Martins; Claudia Renata dos Santos Barros; David Schlesinger; Debora Botequio Moretti; Dimas Tadeu Covas; Elaine Cristina Marqueze; Elaine Vieira Santos; Evandra Strazza Rodrigues; Heidge Fukumasu; Jayme Augusto de Souza-Neto; José Salvatore Leister Patané; Luiz Alcantara; Luiz Lehmann Coutinho; Maria Carolina Elias; Maurício Lacerda Nogueira; Rafael dos Santos Bezerra; Raul Machado Neto; Rejane Maria Tommasini Grotto; Ricardo Haddad; Sandra Coccuzzo Sampaio Vessoni; Simone Kashima; Svetoslav Nanev Slavov; Vincent Louis Viala |
| EPI_ISL_776750, EPI_ISL_776761, EPI_ISL_792101, EPI_ISL_833152, EPI_ISL_861625, EPI_ISL_861626 | Instituto Adolfo Lutz - Central | Instituto Adolfo Lutz, Interdiciplinary Procedures Center, Strategic Laboratory | Claudia Regina Gonçalves; Claudio Tavares Sacchi; Erica Valessa Ramos Gomes; Karoline Rodrigues Campos |
| EPI_ISL_2691193 | Instituto Adolfo Lutz - Reginal de Sao José do Rio Preto | Instituto Adolfo Lutz, Interdiciplinary Procedures Center, Strategic Laboratory | Caio Vinicius Dias Lopes; Claudia Regina Gonçalves; Claudio Tavares Sacchi; Erica Valessa Ramos Gomes; Karoline Rodrigues Campos; Leonardo Jose Tadeu de Araujo |
| EPI_ISL_2691115 | Instituto Adolfo Lutz - Regional de Bauru | Instituto Adolfo Lutz, Interdiciplinary Procedures Center, Strategic Laboratory | Caio Vinicius Dias Lopes; Claudia Regina Gonçalves; Claudio Tavares Sacchi; Erica Valessa Ramos Gomes; Karoline Rodrigues Campos; Leonardo Jose Tadeu de Araujo |
| EPI_ISL_2614532 | Instituto Adolfo Lutz - Regional de Campinas | Instituto Adolfo Lutz, Interdiciplinary Procedures Center, Strategic Laboratory | Caio Vinicius Dias Lopes; Claudia Regina Gonçalves; Claudio Tavares Sacchi; Erica Valessa Ramos Gomes; Karoline Rodrigues Campos; Leonardo Jose Tadeu de Araujo |
| EPI_ISL_4056276 | Instituto Adolfo Lutz - Regional de Santo Andre | Instituto Adolfo Lutz, Interdiciplinary Procedures Center, Strategic Laboratory | Caio Vinicius Dias Lopes; Claudia Regina Gonçalves; Claudio Tavares Sacchi; Karoline Rodrigues Campos; Leonardo Tadeu de Araujo; Marlon Benedito Nascimento Santos |
| EPI_ISL_984246, EPI_ISL_2756432, EPI_ISL_2756445, EPI_ISL_2756480 | Instituto Adolfo Lutz Central | Instituto Adolfo Lutz, Interdiciplinary Procedures Center, Strategic Laboratory | Caio Vinicius Dias Lopes; Claudia Regina Gonçalves; Claudio Tavares Sacchi; Erica Valessa Ramos Gomes; Karoline Rodrigues Campos; Leonardo Jose Tadeu de Araujo |
| EPI_ISL_3418671 | Instituto Carlos Chagas - ICC, FIOCRUZ Parana | Instituto Carlos Chagas - ICC, FIOCRUZ Parana | A.A.; A.M.; A.R.; Aguiar; Albrecht, L.; Alves; Avila; Balsanelli, E.; Becker, G.; Blanes, L.; Dallagiovanna, B.; Debur; E.M.; F.K.; F.O.; Faoro, H.; Graef, T.; H.G.; I.N.; L.G.; L.R.; M.M.; M.O.; Marchini; Md.C.; Morello; Nardeli; Oliveira; P.C.; Passetti, F.; Pedrosa; Resende; Riediger; S.C.; Schemberger; Suzukawa; V.A.; Zanette, D.; de Baura; de Souza; dos Santos |
| EPI_ISL_1678584, EPI_ISL_1690551, EPI_ISL_1694627, EPI_ISL_1694631, EPI_ISL_1694632 | Instituto Estadual do Cérebro Paulo Niemayer (IECPN) | Laboratório de Imunofarmacologia | Thiago Moreno Lopes Souza |
| EPI_ISL_492048 | Instituto de Biologia do Exército | Laboratório Metabolismo Macromolecular FirminoTorres de Castro, Instituto de Biofísica Carlos Chagas Filho, Universidade Federal do Rio de Janeiro | Aline Rosa Vianna de Souza; Bianca Catarina Azevedo Cabral; Caleb GM Santos; Clarissa Damaso; Elizabeth Valentin; Marcio da Costa Cipitelli; Marcos Dornelas-Ribeiro; Nádia Vaez Gonçalves da Cruz; Rodrigo Soares de Moura Neto; Rosane Silva; Tatiana LS Nogueira; Virginia Sara Grancieri do Amaral |
| EPI_ISL_3824342, EPI_ISL_3824344 | Instituto de Biotecnologia - UNESP-Botucatu-SP | Instituto de Biotecnologia - UNESP-Botucatu-SP | Cecília Artico Banho; Cíntia Bittar; Fábio Sossai Possebon; Guilherme Campos; Helena Lage Ferreira; Jorge A. Petroli Marchesi; João Pessoa Araújo Jr.; Leila Sabrina Ullmann; Lívia Sacchetto; Maisa C. Pereira Parra; Marília Moraes; Maurício L. Nogueira; Paula Rahal; Paulo Inacio da Costa |
| EPI_ISL_1054996 | Instituto de Diagnostico y Referencia Epidemiologicos INDRE_RNLSP | Instituto de Diagnostico y Referencia Epidemiologicos (INDRE) | Abril Rodriguez-Maldonado; Adnan Araiza-Rodriguez; Claudia Wong-Arambula; David Fragoso-Fonseca; Ernesto Ramirez-Gonzalez.; Fabiola Garces-Ayala; Gisela Barrera-Badillo; Irma Lopez-Martinez; Lucia Hernandez-Rivas; Mayra Jimenez-Morales; Nancy Munoz-Hernandez; Natividad Cruz-Ortiz; Sergio Rangel-Guerrero; Tatiana Nunez-Garcia |
| EPI_ISL_2691148 | Intituto Adolfo Lutz - Regional de Sorocaba | Instituto Adolfo Lutz, Interdiciplinary Procedures Center, Strategic Laboratory | Caio Vinicius Dias Lopes; Claudia Regina Gonçalves; Claudio Tavares Sacchi; Erica Valessa Ramos Gomes; Karoline Rodrigues Campos; Leonardo Jose Tadeu de Araujo |
| EPI_ISL_837287, EPI_ISL_1039371 | Istituto Zooprofilattico Sperimentale del | TIGEM | Andrea Ballabio; Anna Manfredi; Antonio Grimaldi; Antonio Limone; Biancamaria Pierri; Chiara Colantuono; Davide Cacchiarelli.; Denise Di Concilio; Francesco Panariello; Lucio Di Filippo; Marcello Salvi; Maria Concetta Cuomo; Patrizia Annunziata; Pellegrino Cerino; Valentina Bouche |

| | | | |
|---|---|---|---|
| | Mezzogiorno | | |
| EPI_ISL_1056681 | Istituto Zooprofilattico Sperimentale del Mezzogiorno - IZSM | Telethon Institute of Genetics and Medicine - TIGEM | Andrea Ballabio; Anna Manfredi; Antonio Grimaldi; Antonio Limone; Biancamaria Pierri; Chiara Colantuono; Davide Cacchiarelli.; Denise Di Concilio; Francesco Panariello; Lucio Di Filippo; Marcello Salvi; Maria Concetta Cuomo; Patrizia Annunziata; Pellegrino Cerino; Valentina Bouche |
| EPI_ISL_3218210, EPI_ISL_3218226, EPI_ISL_3218228, EPI_ISL_3218229, EPI_ISL_3218251, EPI_ISL_3218259, EPI_ISL_3218263, EPI_ISL_3218266 | | | |
| see above | LABCOVID_HCPA | LABRESIS_HCPA | Barth AL; Martins AF; Monteiro F; Volpato F; Wink PL; Zavascki AP |
| EPI_ISL_2345896 | LABORATORIO LOCAL DE ITAPECERICA DA SERRA | Instituto Butantan / FZEA-USP-Pirassununga | Antonio Jorge Martins; Claudia Renata dos Santos Barros; David Schlesinger; Debora Botequio Moretti; Dimas Tadeu Covas; Elaine Cristina Marqueze; Elaine Vieira Santos; Evandra Strazza Rodrigues; Heidge Fukumasu; Jayme Augusto de Souza-Neto; José Salvatore Leister Patané; Luiz Alcantara; Luiz Lehmann Coutinho; Maria Carolina Elias; Maurício Lacerda Nogueira; Rafael dos Santos Bezerra; Raul Machado Neto; Rejane Maria Tommasini Grotto; Ricardo Haddad; Sandra Coccuzzo Sampaio Vessoni; Simone Kashima; Svetoslav Nanev Slavov; Vincent Louis Viala |
| EPI_ISL_2493702 | LABORATORIO MUNICIPAL DE ANALISES CLINICAS DE ITANHAEM | Instituto Butantan | Antonio Jorge Martins; Claudia Renata dos Santos Barros; David Schlesinger; Debora Botequio Moretti; Dimas Tadeu Covas; Elaine Cristina Marqueze; Elaine Vieira Santos; Evandra Strazza Rodrigues; Heidge Fukumasu; Jayme Augusto de Souza-Neto; José Salvatore Leister Patané; Luiz Alcantara; Luiz Lehmann Coutinho; Maria Carolina Elias; Maurício Lacerda Nogueira; Rafael dos Santos Bezerra; Raul Machado Neto; Rejane Maria Tommasini Grotto; Ricardo Haddad; Sandra Coccuzzo Sampaio Vessoni; Simone Kashima; Svetoslav Nanev Slavov; Vincent Louis Viala |
| EPI_ISL_2494154 | LABORATORIO MUNICIPAL DE PIRACICABA | Instituto Butantan | Antonio Jorge Martins; Claudia Renata dos Santos Barros; David Schlesinger; Debora Botequio Moretti; Dimas Tadeu Covas; Elaine Cristina Marqueze; Elaine Vieira Santos; Evandra Strazza Rodrigues; Heidge Fukumasu; Jayme Augusto de Souza-Neto; José Salvatore Leister Patané; Luiz Alcantara; Luiz Lehmann Coutinho; Maria Carolina Elias; Maurício Lacerda Nogueira; Rafael dos Santos Bezerra; Raul Machado Neto; Rejane Maria Tommasini Grotto; Ricardo Haddad; Sandra Coccuzzo Sampaio Vessoni; Simone Kashima; Svetoslav Nanev Slavov; Vincent Louis Viala |
| EPI_ISL_1795104, EPI_ISL_1795105, EPI_ISL_2344553, EPI_ISL_2344554, EPI_ISL_2345267 | LABORATORIO MUNICIPAL DE PIRACICABA | Instituto Butantan / ESALQ-Piracicaba | Antonio Jorge Martins; Bianca Cechetto Carlos. Mendelics: Bibiana Santos; Claudia Renata dos Santos Barros; David Schlesinger; David Schlesinger. Hemocentro Ribeirão Preto: Simone Kashima; Debora Botequio Moretti; Debora Botequio Moretti. Centro de Genômica Funcional da ESALQ: Luiz Lehmann Coutinho; Dimas Tadeu Covas; Elaine Cristina Marqueze; Elaine Vieira Santos; Elaine Vieira dos Santos; Elisangela Chicaroni Mattos; Erika Freitas; Evandra Strazza Rodrigues; Felipe Allan da Silva da Costa; Flavia Aburjaile; Guilherme Targino Valente; Heidge Fukumasu; Heidge Fukumasu. USP-Botucatu: Rejane Maria Tommasini Grotto; Instituto Butantan: Alexander Roberto Precioso; Jayme A. Souza-Neto; Jayme Augusto de Souza-Neto; Jessika Cristina Chagas Lesbon; José Salvatore Leister Patané; João Paulo Kitajima; Luiz Alcantara; Luiz Carlos Junior de Alcantara; Luiz Lehmann Coutinho; Maria Carolina Elias; Marta Giovanetti; Maurício Lacerda Nogueira; Patricia Akemi Assato; Rafael dos Santos Bezerra; Raquel de Lello Rocha Campos Cassano. NGS Soluções Genômicas: Pilar Drummond Sampaio Corrêa Mariani. FZEA-USP Pirassununga: Mirele Daiana Poleti; Raul Machado Neto; Rejane Maria Tommasini Grotto; Ricardo Augusto Brassaloti; Ricardo Haddad; Rodrigo Tocantins Calado.; Sandra Coccuzzo Sampaio; Sandra Coccuzzo Sampaio Vessoni; Simone Kashima; Svetoslav Nanev Slavov; Vagner Fonseca; Vincent Louis Viala |
| EPI_ISL_943989, EPI_ISL_985315 | LACEN do Estado de Goias | Instituto Adolfo Lutz, Interdiciplinary Procedures Center, Strategic Laboratory | Claudia Regina Gonçalves; Claudio Tavares Sacchi; Erica Valessa Ramos Gomes; Karoline Rodrigues Campos |
| EPI_ISL_3671916 | LACEN do Estado do Mato Grosso do Sul | Instituto Adolfo Lutz, Interdiciplinary Procedures Center, Strategic Laboratory | Caio Vinicius Dias Lopes; Claudia Regina Gonçalves; Claudio Tavares Sacchi; Karoline Rodrigues Campos; Leonardo Tadeu de Araujo; Marlon Benedito Nascomento Santos |
| EPI_ISL_1139060, EPI_ISL_1139068, EPI_ISL_1201883 | LACEN do Mato Grosso do Sul | Instituto Adolfo Lutz, Interdiciplinary Procedures Center, Strategic Laboratory | Caio Vinicius Dias Lopes; Claudia Regina Gonçalves; Claudio Tavares Sacchi; Erica Valessa Ramos Gomes; Karoline Rodrigues Campos |
| EPI_ISL_3060267 | LACEN/PE | WallauLab on behalf of Fiocruz COVID-19 Genomic Surveillance Network | Alexandre Freitas da Silva; Cassia Docena; Constância Flávia Junqueira Ayres; Filipe Zimmer Dezordi; Gabriel Luz Wallau; Gustavo Barbosa de Lima; Lais Ceschini Machado; Lilian Caroliny Amorim Silva; Marcelo Henrique dos Santos Paiva; Matheus Filgueira Bezerra; Sinval Pinto Brandão Filho |
| EPI_ISL_4488002 | LACEN/PI | WallauLab on behalf of Fiocruz COVID-19 Genomic Surveillance Network | Adelino Soares Lima Neto; Alexandre Freitas da Silva; Antônio Marinho; Cassia Docena; Constância Flávia Junqueira Ayres; Filipe Zimmer Dezordi; Gabriel Luz Wallau; Gustavo Barbosa de Lima; Hellen de Oliveira Amaral; Jacenir Reis dos Santos Mallet; Joana Carolina Viana Lima; Lais Ceschini Machado; Leandro de Mattos; Lilian Caroliny Amorim Silva; Marcela de Lacerda Valença Queiroz; Marcelo Adriano da Cunha e Silva Vieira; Marcelo Henrique dos Santos Paiva; Matheus Filgueira Bezerra; Sinval Pinto Brandão Filho; Túlio de Lima Campos; Vladimir Costa Silva; Walterlene de Carvalho Goçalves |
| EPI_ISL_861896, EPI_ISL_861900, EPI_ISL_861902, EPI_ISL_861905, EPI_ISL_861906, EPI_ISL_861912, EPI_ISL_861913 | | | |
| see above | LATE - Laboratório de Técnicas Especiais - Hospital Israelita Albert Einstein | LATE - Laboratório de Técnicas Especiais - Hospital Israelita Albert Einstein | Ana Paula Moreira Salles; Deyvid Amgarten; Fernanda de Mello Malta; João Renato Rebello Pinho; Pedro Henrique Sebe Rodrigues; Raquel Riyuzo |
| EPI_ISL_429718, EPI_ISL_429758 | Laboratoire National de Sante, Microbiology, Virology | Laboratoire National de Sante, Microbiology, Epidemiology and Microbial Genomics | Anke Wienecke-Baldacchino; Ardashel Latsuzbaia; Catherine Ragimbeau; Guillaume Fournier; Jessica Tapp; Joel Mossong; Tamir Abdelrahman; Trung Nguyen Nguyen |
| EPI_ISL_740404, EPI_ISL_744681 | Laboratoire national de santé, Microbiology, Virology | Laboratoire national de santé, Microbiology, Microbial Genomics Platform | Anke Wienecke-Baldacchino; Catherine Ragimbeau; Fatu Djabi; Jessica Tapp; Tamir Abdelrahman |
| EPI_ISL_4080849 | Laboratorio Central de Saude Publica do Estado de Goias (LACEN/GO) | Laboratory of Respiratory Viruses and Measles, Oswaldo Cruz Institute, FIOCRUZ | Agatha Cristinne Prudencio; Alice Sampaio Rocha; Ana Carolina Mendonca; Ana Flavia Mendonça; Anna Carolina Paixao; Carmen Helena Ramos; Cassiane Casanova; Elisa Cavalcante Pereira; Fernando Motta; Flavia Pereira Amorim da Silva; Ighor Leonardo Arantes Gomes; Luciana Appolinario; Luiz Augusto Pereira; Marilda Siqueira on behalf of the Fiocruz COVID-19 Genomic Surveillance Network; Paola Resende; Rafael Souza Guedes; Renata Serrano Lopes; Taina Venas; Vinicius Lemes da Silva |
| EPI_ISL_2645877 | Laboratorio Central de Saude Publica do Estado do Para (LACEN/PA) | Laboratory of Respiratory Viruses and Measles, Oswaldo Cruz Institute, FIOCRUZ | Alice Sampaio Rocha; Ana Carolina Mendonca; Anna Carolina Paixao; Elisa Cavalcante Pereira; Fernando Motta; Luciana Appolinario; Marilda Siqueira on behalf of the Fiocruz COVID-19 Genomic Surveillance Network; Paola Resende; Renata Serrano Lopes; Taina Venas; Valnete Andrade |
| EPI_ISL_2101689, EPI_ISL_3245237 | Laboratorio Central Noel Nutels | Bioinformatics Laboratory / LNCC | Alessandra P Lamarca; Alexandra L Gerber; Amilcar Tanuri; Ana Paula de C Guimaraes; Ana Tereza R Vasconcelos; Andrea Cony Cavalcanti; Caio Luiz Pereira Ribeiro; Cassia Alves; Cintia Policarpo; Claudia Maria Braga de Mello; Cristiane Gomes da Silva; Diana Mariani; Douglas Terra Machado; Erica Ramos dos Santos Nascimento; Fernanda Leitao dos Santos; Flavio Dias da Silva; Gleidson da Silva de Oliveira; Leandro Magalhaes de Souza; Liliane Cavalcante; Luiz G P de Almeida; Marcio Henrique de Oliveira Garcia; Mario Sergio Ribeiro; Ricardo Jose Barbosa Salviano; Ronaldo da Silva F Jr; Silvia Carvalho |
| EPI_ISL_2536303, EPI_ISL_2536316, EPI_ISL_2731457 | Laboratorio Central de Saude Publica do Estado da Paraiba (LACEN-PB) | Laboratory of Respiratory Viruses and Measles, Oswaldo Cruz Institute, FIOCRUZ | Alice Sampaio Rocha; Ana Carolina Mendonca; Anna Carolina Paixao; Dalane Loudal Florentino Teixeira; Elisa Cavalcante Pereira; Fernando Motta; Joao Felipe Bezerra; Luciana Appolinario; Marilda Siqueira on behalf of the Fiocruz COVID-19 Genomic Surveillance Network; Paola Resende; Renata Serrano Lopes; Taina Venas |
| EPI_ISL_2645660, EPI_ISL_2645663, EPI_ISL_2645668, EPI_ISL_2645674, EPI_ISL_2645680, EPI_ISL_2645692 | Laboratorio Central de Saude Publica do Estado de Alagoas (LACEN/AL) | Laboratory of Respiratory Viruses and Measles, Oswaldo Cruz Institute, FIOCRUZ | Alice Sampaio Rocha; Ana Carolina Mendonca; Anderson Brandao Leite; Anna Carolina Paixao; Elisa Cavalcante Pereira; Fernando Motta; Luciana Appolinario; Marilda Siqueira on behalf of the Fiocruz COVID-19 Genomic Surveillance Network; Paola Resende; Renata Serrano Lopes; Taina Venas |
| EPI_ISL_2660462, EPI_ISL_2660491, EPI_ISL_2660542 | Laboratorio Central de Saude Publica do Estado de Minas Gerais (LACEN/MG) | Laboratory of Respiratory Viruses and Measles, Oswaldo Cruz Institute, FIOCRUZ | Alice Sampaio Rocha; Ana Carolina Mendonca; Andre Felipe Leal Bernardes; Anna Carolina Paixao; Elisa Cavalcante Pereira; Fernando Motta; Luciana Appolinario; Marilda Siqueira on behalf of the Fiocruz COVID-19 Genomic Surveillance Network; Paola Resende; Renata Serrano Lopes; Taina Venas |
| EPI_ISL_1534003 | Laboratorio Central de Saude Publica do Estado de Santa Catarina (LACEN-SC) | Laboratory of Respiratory Viruses and Measles, Oswaldo Cruz Institute, FIOCRUZ | Alice Sampaio Rocha; Ana Carolina Mendonca; Anna Carolina Paixao; Darcita Buerger Rovaris; Fernando Motta; Luciana Appolinario; Marilda Siqueira on behalf of the Fiocruz COVID-19 Genomic Surveillance Network; Paola Resende; Renata Serrano Lopes; Sandra Bianchini Fernandes |
| EPI_ISL_4563059, EPI_ISL_4563061 | Laboratorio Central de Saude Publica do Estado de Santa Catarina (LACEN/SC) | Laboratory of Respiratory Viruses and Measles, Oswaldo Cruz Institute, FIOCRUZ | Alice Sampaio Rocha; Ana Carolina Mendonca; Anna Carolina Paixao; Darcita Buerger Rovaris; Elisa Cavalcante Pereira; Fernando Motta; Luciana Appolinario; Marilda Siqueira on behalf of the Fiocruz COVID-19 Genomic Surveillance Network; Paola Resende; Renata Serrano Lopes; Sandra Bianchini Fernandes; Taina Venas |
| EPI_ISL_2645521, EPI_ISL_2645529, EPI_ISL_2645544, EPI_ISL_2645571, EPI_ISL_3061879 | Laboratorio Central de Saude Publica do Estado do Espirito Santo (LACEN/ES) | Laboratory of Respiratory Viruses and Measles, Oswaldo Cruz Institute, FIOCRUZ | Alice Sampaio Rocha; Ana Carolina Mendonca; Anna Carolina Paixao; Eilisa Cavalcante Pereira; Fernando Motta; Luciana Appolinario; Marilda Siqueira on behalf of the Fiocruz COVID-19 Genomic Surveillance Network; Paola Resende; Renata Serrano Lopes; Rodrigo Ribeiro Rodrigues; Taina Venas |
| EPI_ISL_2731458, EPI_ISL_3061892, EPI_ISL_3061893, EPI_ISL_3243105, EPI_ISL_3243106, EPI_ISL_3243107, EPI_ISL_3243108, EPI_ISL_4061296, EPI_ISL_4880350 | | | |
| see above | Laboratorio Central de Saude Publica do Estado do Parana (LACEN/PR) | Laboratory of Respiratory Viruses and Measles, Oswaldo Cruz Institute, FIOCRUZ | Agatha Soares; Alice Sampaio Rocha; Ana Carolina Mendonca; Anna Carolina Paixao; Elisa Cavalcante Pereira; Fernando Motta; Ighor Leonardo Arantes; Irina Riediger; Luciana Appolinario; Marilda Siqueira on behalf of the Fiocruz COVID-19 Genomic Surveillance Network; Paola Resende; Renata Serrano Lopes; Taina Venas |
| EPI_ISL_1533979, EPI_ISL_2274108, EPI_ISL_2603449, EPI_ISL_2603549, EPI_ISL_2603584, EPI_ISL_2661821, EPI_ISL_2661837, EPI_ISL_2661844, EPI_ISL_2661845, EPI_ISL_2661847, EPI_ISL_2661854, EPI_ISL_2661855, EPI_ISL_2661856 | | | |

| | | | |
|---|---|---|---|
| see above | Laboratorio Central de Saude Publica do Estado do Rio Grande do Sul (LACEN-RS) | Laboratory of Respiratory Viruses and Measles, Oswaldo Cruz Institute, FIOCRUZ | Alice Sampaio Rocha; Ana Carolina Mendonca; Anderson Brandao Leite; Anna Carolina Paixao; Elisa Cavalcante Pereira; Fernando Motta; Luciana Appolinario; Marilda Siqueira on behalf of the Fiocruz COVID-19 Genomic Surveillance Network; Paola Resende; Renata Serrano Lopes; Richard Salvato; Taina Venas; Tatiana Schaffer Gregianini |
| EPI_ISL_3048758, EPI_ISL_3048759, EPI_ISL_3048773, EPI_ISL_3048774 | Laboratorio Central de Saude Publica do Estado do Rio Grande do Sul (LACEN-RS) | Laboratório de Biologia Molecular da Universidade Federal de Ciências da Saúde de Porto Alegre | Adriana Seixas; Ana B. G. Veiga; Ana Paula Muterle Varela; Fabiana Quoos Mayer; Fernando Hayashi Sant'Anna; Janira Prichula; Letícia Garay Martins; Richard Steiner Salvato; Tatiana Schäffer Gregianini |
| EPI_ISL_2970372 | Laboratorio de Biologia Molecular de Flavivirus, Instituto Oswaldo Cruz | Laboratorio de Biologia Molecular de Flavivirus, Instituto Oswaldo Cruz | A.A.; Bonaldo; Brasil, P.; Damasceno, L.; Dias, B.; I.P.; L.M.; M.C.; Pelajo, M.; Raphael; Ribeiro; Santos |
| EPI_ISL_811148, EPI_ISL_811149, EPI_ISL_1034304, EPI_ISL_1034305, EPI_ISL_1034306, EPI_ISL_1068078, EPI_ISL_1068085, EPI_ISL_1068118, EPI_ISL_1068119, EPI_ISL_1068164, EPI_ISL_1068165, EPI_ISL_1068171, EPI_ISL_1068174, EPI_ISL_1068176, EPI_ISL_1068177, EPI_ISL_1068178, EPI_ISL_1068179, EPI_ISL_1068180, EPI_ISL_1068181, EPI_ISL_1068183, EPI_ISL_1068184, EPI_ISL_1068185, EPI_ISL_1068186, EPI_ISL_1068187, EPI_ISL_1068188, EPI_ISL_1068191, EPI_ISL_1068193, EPI_ISL_1068194, EPI_ISL_1068204, EPI_ISL_1068219, EPI_ISL_1068238, EPI_ISL_1068239, EPI_ISL_1114151 | | | |
| see above | Laboratorio de Ecologia de Doencas Transmissiveis na Amazonia, Instituto Leonidas e Maria Deane - Fiocruz Amazonia | Laboratorio de Ecologia de Doencas Transmissiveis na Amazonia, Instituto Leonidas e Maria Deane - Fiocruz Amazonia | André Corado; Debora Duarte; Felipe Naveca on behalf of the Fiocruz COVID-19 Genomic Surveillance Network; Fernanda Nascimento; George Silva; Karina Pessoa; Luciana Gonçalves; Maria Júlia Brandão; Matilde Mejía; Michele Jesus; Valdinete Nascimento; Victor Souza; Ágatha Costa |
| EPI_ISL_2544883, EPI_ISL_3049016 | Laboratorio de Pesquisa em Virologia, FAMERP, SJRP | Laboratorio de Pesquisa em Virologia, FAMERP, SJRP | Cecília Artico Banho; Cíntia Bittar; Fábio Sossai Possebon; Guilherme Campos; Helena Lage Ferreira; Jorge A. Petroli Marchesi; João Pessoa Araújo Jr.; Leila Sabrina Ullmann; Lívia Sacchetto; Maisa C. Pereira Parra; Marília Moraes; Maurício L. Nogueira.; Paula Rahal; Paulo Inacio da Costa |
| EPI_ISL_623130, EPI_ISL_623140, EPI_ISL_623142 | Laboratorio de Virologia Molecular / UFRJ | Bioinformatics Laboratory / LNCC | Alexandra L Gerber; Amilcar Tanuri; Ana Paula de C Guimarães; Ana Tereza R de Vasconcelos; Carolina M Voloch; Covid19-UFRJ Workgroup; Cynthia C Cardoso; Diana Mariani; Luiz G P de Almeida; Luís Cristóvão Pôrto; Orlando C. Ferreira; Otavio J. Brustolini; Renato S Aguiar; Ronaldo S Francisco Jr; Terezinha M P P Castiñeiras |
| EPI_ISL_910335 | Laboratory for Respiratory Viruses, Cantacuzino National Military-Medical Institute for Research and Development | Cantacuzino Institute Virology | Luiza Ustea; Mihaela Lazar; Nicoleta Paraschiv |
| EPI_ISL_2196251, EPI_ISL_2196361, EPI_ISL_2614111, EPI_ISL_2614113, EPI_ISL_2614114, EPI_ISL_2614117, EPI_ISL_2614137, EPI_ISL_2614159, EPI_ISL_2677181, EPI_ISL_2677207 | | | |
| see above | Laboratory of Respiratory Viruses and Measles, Oswaldo Cruz Institute, FIOCRUZ | Laboratory of Respiratory Viruses and Measles, Oswaldo Cruz Institute, FIOCRUZ | Alice Sampaio Rocha; Ana Carolina Mendonca; Anna Carolina Paixao; Elisa Cavalcante Pereira; Fernando Motta; Luciana Appolinario; Marilda Siqueira on behalf of the Fiocruz COVID-19 Genomic Surveillance Network; Paola Resende; Renata Serrano Lopes; Taina Venas |
| EPI_ISL_2157396, EPI_ISL_2157399, EPI_ISL_2157447 | Laboratório Central de Saude Publica do Estado de Santa Catarina (LACEN/SC) | Laboratory of Respiratory Viruses and Measles, Oswaldo Cruz Institute, FIOCRUZ | Alice Sampaio Rocha; Ana Carolina Mendonca; Anna Carolina Paixao; Darcita Buerger Rovaris; Elisa Cavalcante Pereira; Fernando Motta; Luciana Appolinario; Marilda Siqueira on behalf of the Fiocruz COVID-19 Genomic Surveillance Network; Paola Resende; Renata Serrano Lopes; Sandra Bianchini Fernandes; Taina Venas |
| EPI_ISL_2241571 | Laboratório Central de Saúde Pública da Paraiba | Coordenação Geral de Laboratórios de Saúde Pública (CGLAB/DAEVS/SVS/MS) | Vagner Fonseca; et al. |
| EPI_ISL_2249398 | Laboratório Central de Saúde Pública de Santa Catarina | Coordenação Geral de Laboratórios de Saúde Pública (CGLAB/DAEVS/SVS/MS) | Vagner Fonseca; et al. |
| EPI_ISL_2248780 | Laboratório Central de Saúde Pública do Acre | Coordenação Geral de Laboratórios de Saúde Pública (CGLAB/DAEVS/SVS/MS) | Vagner Fonseca; et al. |
| EPI_ISL_2298824 | Laboratório Central de Saúde Pública do Amapá | Coordenação Geral de Laboratórios de Saúde Pública (CGLAB/DAEVS/SVS/MS) | Vagner Fonseca; et al. |
| EPI_ISL_2298745 | Laboratório Central de Saúde Pública do Ceará | Coordenação Geral de Laboratórios de Saúde Pública (CGLAB/DAEVS/SVS/MS) | Vagner Fonseca; et al. |
| EPI_ISL_1239118 | Laboratório Central de Saúde Pública do Espírito Santo | Coordenação Geral de Laboratórios de Saúde Pública (CGLAB) | ; Vagner Fonseca et al |
| EPI_ISL_2293013 | Laboratório Central de Saúde Pública do Espírito Santo | Coordenação Geral de Laboratórios de Saúde Pública (CGLAB/DAEVS/SVS/MS) | Vagner Fonseca; et al. |
| EPI_ISL_541374 | Laboratório Central de Saúde Pública do Estado de Sergipe (LACEN-SE) | Laboratory of Respiratory Viruses and Measles, Oswaldo Cruz Institute, FIOCRUZ | Ana Carolina Mendonça; Anna Carolina Paixão; Clioma Santos; Fernando Motta; Jonathan Lopes; Luciana Appolinario; Marilda Siqueira on behalf of the Fiocruz COVID-19 Genomic Surveillance Network; Paola Resende |
| EPI_ISL_792647 | Laboratório Central de Saúde Pública do Estado do Paraná (LACEN-PR) | Laboratory of Respiratory Viruses and Measles, Oswaldo Cruz Institute, FIOCRUZ | Ana Carolina Mendonca; Anna Carolina Paixao; Fernando Motta; Irina Nastassja Riediger; Luciana Appolinario; Maria do Carmo Debur; Marilda Siqueira on behalf of the Fiocruz COVID-19 Genomic Surveillance Network; Paola Resende |
| EPI_ISL_2731631 | Laboratório de Biotecnologia Aplicada (LBA) - Laboratório de Biologia Molecular - Hospital das Clínicas, Faculdade de Medicina de Botucatu, Departamento de Bioprocessos e Biotecnologia - Faculdade de Ciências Agronômicas, UNESP – Botucatu/SP | Laboratory of Respiratory Viruses and Measles, Oswaldo Cruz Institute, FIOCRUZ | Alice Sampaio Rocha; Ana Carolina Mendonca; Anna Carolina Paixao; Elisa Cavalcante Pereira; Felipe Allan da Silva Costa; Fernando Motta; Jayme Augusto de Souza Neto; Leonardo Nazario de Moraes; Luciana Appolinario; Marilda Siqueira on behalf of the Fiocruz COVID-19 Genomic Surveillance Network; Paola Resende; Patricia Akemi Assato; Rejane Maria Tommasini; Renata Serrano Lopes; Taina Venas |
| EPI_ISL_1464671 | Laboratório de Virologia - UNIFESP | Laboratory of Respiratory Viruses and Measles, Oswaldo Cruz Institute, FIOCRUZ | Alice Sampaio Rocha; Ana Carolina Mendonca; Anna Carolina Paixao; Fernando Motta; Luciana Appolinario; Marilda Siqueira on behalf of the Fiocruz COVID-19 Genomic Surveillance Network; Nancy Belei; Paola Resende; Renata Serrano Lopes |
| EPI_ISL_2196249, EPI_ISL_2196250, EPI_ISL_2196357, EPI_ISL_2196360, EPI_ISL_2443703, EPI_ISL_2536228, EPI_ISL_2677131, EPI_ISL_2677132, EPI_ISL_2677313, EPI_ISL_3061900, EPI_ISL_3061901, EPI_ISL_3061902 | | | |
| see above | Labortorio Central de Saude Publica do Estado de Santa Catarina (LACEN/SC) | Laboratory of Respiratory Viruses and Measles, Oswaldo Cruz Institute, FIOCRUZ | Alice Sampaio Rocha; Ana Carolina Mendonca; Anna Carolina Paixao; Darcita Buerger Rovaris; Elisa Cavalcante Pereira; Fernando Motta; Luciana Appolinario; Marilda Siqueira on behalf of the Fiocruz COVID-19 Genomic Surveillance Network; Paola Resende; Renata Serrano Lopes; Sandra Bianchini Fernandes; Taina Venas |
| EPI_ISL_2443646 | Labortorio Central de | Laboratory of | Alice Sampaio Rocha; Ana Carolina Mendonca; Andrea Cony Cavalcanti; Anna Carolina Paixao; Elisa Cavalcante Pereira; Fernando Motta; Luciana Appolinario; Marilda Siqueira on behalf of the Fiocruz COVID-19 Genomic Surveillance Network; Paola Resende; Renata Serrano Lopes; Taina Venas |

| | | | |
|---|---|---|---|
| | Saude Publica do Estado do Rio de Janeiro (LACEN/RJ) | Respiratory Viruses and Measles, Oswaldo Cruz Institute, FIOCRUZ | |
| EPI_ISL_2645509, EPI_ISL_2645511, EPI_ISL_3190409 | Labortorio Central de Saude Publica do Estado do Tocantins (LACEN/TO) | Laboratory of Respiratory Viruses and Measles, Oswaldo Cruz Institute, FIOCRUZ | Agatha Soares; Alice Sampaio Rocha; Ana Carolina Mendonca; Anna Carolina Paixao; Elisa Cavalcante Pereira; Fernando Motta; Ighor Arantes; Jucimaria Dantas Galvao; Luciana Appolinario; Marilda Siqueira on behalf of the Fiocruz COVID-19 Genomic Surveillance Network; Paola Resende; Renata Serrano Lopes; Taina Venas |
| EPI_ISL_2196252, EPI_ISL_2196362, EPI_ISL_2603462, EPI_ISL_2603463, EPI_ISL_2603544 | Lboratorio Central de Saude Publica do Estado do Parana (LACEN/PR) | Laboratory of Respiratory Viruses and Measles, Oswaldo Cruz Institute, FIOCRUZ | Alice Sampaio Rocha; Ana Carolina Mendonca; Anna Carolina Paixao; Elisa Cavalcante Pereira; Fernando Motta; Irina Riediger; Luciana Appolinario; Marilda Siqueira on behalf of the Fiocruz COVID-19 Genomic Surveillance Network; Paola Resende; Renata Serrano Lopes; Taina Venas |
| EPI_ISL_3447543 | NUPIT/UFPE | WallauLab on behalf of Fiocruz COVID-19 Genomic Surveillance Network | Alexandre Freitas da Silva; Cassia Docena; Constância Flávia Junqueira Ayres; Filipe Zimmer Dezordi; Gabriel Luz Wallau; Gustavo Barbosa de Lima; Lais Ceschini Machado; Lilian Caroliny Amorim Silva; Maira Galdino da Rocha Pitta; Marcelo Henrique dos Santos Paiva; Matheus Filgueira Bezerra; Michelly Cristiny Pereira; Rômulo Pessoa e Silva; Sinval Pinto Brandão Filho |
| EPI_ISL_416028 | National Influenza Center - Instituto Adolfo Lutz | Instituto Adolfo Lutz, Interdiciplinary Procedures Center, Strategic Laboratory | Adriana Bugno; Adriano Abbud; Carlos Henrique Camargo; Claudia Regina Gonçalves; Claudio Tavares Sacchi; Daniela Bernardes Borges da Silva; Fabiana Cristina Pereira dos Santos; Maria do Carmo Sampaio Tavares Timenetsky; Simone Guadagnucci Morillo; Terezinha Maria de Paiva |
| EPI_ISL_524620 | North West London Pathology, Imperial College Healthcare NHS Trust | Wellcome Sanger Institute for the COVID-19 Genomics UK (COG-UK) consortium | Aileen Rowan; Alison Holmes; Anjna Badhan; Carolina Herrera and Alex Alderton; Cordelia Langford; David K. Jackson; David Muir; Dominic Kwiatkowski; Ewan Harrison; Frankie Bolt; Graham Taylor; Ian Johnston; James Price; John Sillitoe on behalf of the Wellcome Sanger Institute COVID-19 Surveillance Team (http://www.sanger.ac.uk/covid-team); Ling Li; Paul Randell; Roberto Amato; Sonia Goncalves |
| EPI_ISL_1117411, EPI_ISL_1117418 | Nucleo de Pesquisa em Inovacao Terapeutica - UFPE | LABBE, Federal University of Pernambuco | Bruno Sampaio; Heidi Lacerda Alves da Cruz; Maira Galdino da Rocha Pitta; Marco Katzenberger; Marcos da Silveira Regueira Neto; Michelly Cristiny Pereira; Reginaldo Goncalves de Lima Neto; Valdir de Queiroz Balbino; Wilson Jose da Silva Junior |
| EPI_ISL_1181622 | Oswaldo Cruz Foundation, FIOCRUZ - Ceara (Fiocruz-CE) | Laboratory of Respiratory Viruses and Measles, Oswaldo Cruz Institute, FIOCRUZ | Alice Sampaio Rocha; Ana Carolina Mendonca; Anna Carolina Paixao; Fabio Miyajima; Fernando Motta; Joaquim César do Nascimento Sousa Júnior; Luciana Appolinario; Marilda Siqueira on behalf of the Fiocruz COVID-19 Genomic Surveillance Network; Paola Resende; Renata Serrano Lopes; Thais de Oliveira Costa |
| EPI_ISL_572714, EPI_ISL_741057 | Oxford Viromics, NDM, University of Oxford; Oxford University Hospitals; Basingstoke and North Hampshire Hospital | COVID-19 Genomics UK (COG-UK) Consortium | Alex Mobbs; Amy Trebes; Anita Justice; Catrin Moore; Christophe Fraser; David Bonsall; David Buck; Emma Wise; George Macintyre; Jessica Lynch; John Todd; Mariateresa de Cesare; Matilde Mori; Monique Andersson; Nathan Moore; Nick Cortes; Robert Shaw; Stephen Kidd; Tanya Golubchik; Timothy Peto |
| EPI_ISL_2345356, EPI_ISL_2345367, EPI_ISL_2345401 | POLICLINICA HORTOLANDIA | Instituto Butantan / ESALQ-Piracicaba | Antonio Jorge Martins; Claudia Renata dos Santos Barros; David Schlesinger; Debora Botequio Moretti; Dimas Tadeu Covas; Elaine Cristina Marqueze; Elaine Vieira Santos; Evandra Strazza Rodrigues; Heidge Fukumasu; Jayme Augusto de Souza-Neto; José Salvatore Leister Patané; Luiz Alcantara; Luiz Lehmann Coutinho; Maria Carolina Elias; Maurício Lacerda Nogueira; Rafael dos Santos Bezerra; Raul Machado Neto; Rejane Maria Tommasini Grotto; Ricardo Haddad; Sandra Coccuzzo Sampaio Vessoni; Simone Kashima; Svetoslav Nanev Slavov; Vincent Louis Viala |
| EPI_ISL_2663276, EPI_ISL_2663287 | Plataforma de Vigilancia Molecular (PVM) - FIOCRUZ/BA | Plataforma de Vigilancia Molecular (PVM) - FIOCRUZ/BA | Bruno Bezerril Andrade; Camila I. de Oliveira on behalf of the Fiocruz COVID-19 Genomic Surveillance Network.; Clarissa Araújo Gurgel; Leonardo Paiva Farias; Marina Cucco; Ricardo Khouri; Tiago Graf |
| EPI_ISL_2344627, EPI_ISL_2346089 | Prefeirtura de SP | Instituto Butantan | Antonio Jorge Martins; Claudia Renata dos Santos Barros; David Schlesinger; Debora Botequio Moretti; Dimas Tadeu Covas; Elaine Cristina Marqueze; Elaine Vieira Santos; Evandra Strazza Rodrigues; Heidge Fukumasu; Jayme Augusto de Souza-Neto; José Salvatore Leister Patané; Luiz Alcantara; Luiz Lehmann Coutinho; Maria Carolina Elias; Maurício Lacerda Nogueira; Rafael dos Santos Bezerra; Raul Machado Neto; Rejane Maria Tommasini Grotto; Ricardo Haddad; Sandra Coccuzzo Sampaio Vessoni; Simone Kashima; Svetoslav Nanev Slavov; Vincent Louis Viala |
| EPI_ISL_693231 | Pronto Socorro Municipal de Santa Branca | Instituto Adolfo Lutz, Interdiciplinary Procedures Center, Strategic Laboratory | Claudia Regina Gonçalves; Claudio Tavares Sacchi; Erica Valessa Ramos Gomes; Karoline Rodrigues Campos |
| EPI_ISL_577107, EPI_ISL_595206, EPI_ISL_595224 | Quadram Institute Bioscience | COVID-19 Genomics UK (COG-UK) Consortium | Alexander J Trotter; Alison E. Mather; Alp Aydin; Ana P. Tedim; Anastasia Kolyva; Andrew Bell; Andrew J. Page; Claire Stuart; Dave J. Baker; Gemma L. Kay; John Wain; Justin O'Grady; Leonardo de Oliveira Martins; Lizzie Meadows; Maria Diaz; Mark Webber; Muhammed Yasir; Nabil-Fareed Alikhan; Ngozi Elumogo; Nicholas M. Thomson; Rachael Stanley; Rachel Gilroy; Reenesh Prakash; Samir Dervisevic; Samuel Bloomfield; Steven Rudder; Thanh Le-Viet |
| EPI_ISL_693254 | Queensland Health Forensic and Scientific Services | Queensland Health Forensic and Scientific Services | Son Nguyen et al |
| EPI_ISL_585172 | Regional Virus Laboratory, Belfast Health and Social Care Trust | COVID-19 Genomics UK (COG-UK) Consortium | Alison Watt; Ciara Cox; Conall McCaughey; David Simpson; Derek Fairley; James McKenna; Mairead Connor; Susan Feeney; Tanya Curran; Zoltan Molnar |
| EPI_ISL_2734071 | Respiratory Virus Unit, Microbiology Services Colindale, Public Health England | COVID-19 Genomics UK (COG-UK) Consortium | PHE Covid Sequencing Team |
| EPI_ISL_1967019 | SANTA CASA DE PORTO FELIZ | Instituto Butantan / ESALQ-USP (Piracicaba) | Antonio Jorge Martins; Bianca Cechetto Carlos. Mendelics: Bibiana Santos; Claudia Renata dos Santos Barros; Cíntia Bittar; David Schlesinger. Hemocentro Ribeirão Preto: Simone Kashima; Debora Botequio Moretti; Elaine Cristina Marqueze; Elaine Vieira dos Santos; Elisangela Chicaroni Mattos; Erika Freitas; Evandra Strazza Rodrigues; Felipe Allan da Silva da Costa; Flavia Aburjaile; Fábio Sossai Possebon; Guilherme Campos; Guilherme Targino Valente; Heidge Fukumasu. USP-Botucatu: Rejane Maria Tommasini Grotto; Helena Lage Ferreira; Instituto Butantan: Dimas Tadeu Covas; Jardelina de Souza Todao Bernardino; Jayme A. Souza-Neto; Jessika Cristina Chagas Lesbon; Jorge A. Petroli Marchesi; José Salvatore Leister Patané; João Paulo Kitajima; João Pessoa Araújo Jr.; Leila Sabrina Ullmann; Loyze Paola Oliveira de Lima; Luiz Aurelio de Campos Crispin. Centro de Genômica Funcional da ESALQ: Luiz Lehmann Coutinho; Luiz Carlos Junior de Alcantara; Lívia Sacchetto; Maisa C. Pereira Parra; Maria Carolina Elias; Marta Giovanetti; Marília Moraes; Maurício Lacerda Nogueira. Prefeitura de Sao Paulo: Melissa Palmieri.; Patricia Akemi Assato; Paula Rahal; Paulo Inacio da Costa; Rafael dos Santos Bezerra; Raquel de Lello Rocha Campos Cassano. NGS Soluções Genômicas: Pilar Drummond Sampaio Corrêa Mariani. FZEA-USP Pirassununga: Mirele Daiana Poleti; Raul Machado Neto; Ricardo Augusto Brassaloti; Ricardo Haddad; Rodrigo Tocantins Calado. FAMERP-SJRP: Cecília Artico Banho; Sandra Coccuzzo Sampaio; Svetoslav Nanev Slavov; Vagner Fonseca; Vincent Louis Viala |
| EPI_ISL_3102463 | SAO CARLOS DIAGNOSTICO POR IMAGEM | Analytical Competence Molecular Epidemiology Lab/ACME, Oswaldo Cruz Foundation, Ceara (FIOCRUZ CE) | Cleber Furtado Aksenen; Fabio Miyajima; Fernando Braga Stehling; Francisco Eder de Moura Lopes; Jamille Maria Mendes Bezerra; Joaquim César do Nascimento Sousa Junior; Pedro Miguel Carneiro Jeronimo; Suzana Porto Almeida e Lucas Delerino; Thais Ferreira de Oliveira; Thais de Oliveira Costa; Ticiane Cavalcante de Souza; Veridiana Pessoa Miyajima |
| EPI_ISL_2345849 | SECRETARIA DE SAUDE PUBLICA DE PRAIA GRANDE | Instituto Butantan / Mendelics | Antonio Jorge Martins; Claudia Renata dos Santos Barros; David Schlesinger; Debora Botequio Moretti; Dimas Tadeu Covas; Elaine Cristina Marqueze; Elaine Vieira Santos; Evandra Strazza Rodrigues; Heidge Fukumasu; Jayme Augusto de Souza-Neto; José Salvatore Leister Patané; Luiz Alcantara; Luiz Lehmann Coutinho; Maria Carolina Elias; Maurício Lacerda Nogueira; Rafael dos Santos Bezerra; Raul Machado Neto; Rejane Maria Tommasini Grotto; Ricardo Haddad; Sandra Coccuzzo Sampaio Vessoni; Simone Kashima; Svetoslav Nanev Slavov; Vincent Louis Viala |
| EPI_ISL_2493690 | SECRETARIA MUNICIPAL DE SAUDE SOROCABA | Instituto Butantan | Antonio Jorge Martins; Claudia Renata dos Santos Barros; David Schlesinger; Debora Botequio Moretti; Dimas Tadeu Covas; Elaine Cristina Marqueze; Elaine Vieira Santos; Evandra Strazza Rodrigues; Heidge Fukumasu; Jayme Augusto de Souza-Neto; José Salvatore Leister Patané; Luiz Alcantara; Luiz Lehmann Coutinho; Maria Carolina Elias; Maurício Lacerda Nogueira; Rafael dos Santos Bezerra; Raul Machado Neto; Rejane Maria Tommasini Grotto; Ricardo Haddad; Sandra Coccuzzo Sampaio Vessoni; Simone Kashima; Svetoslav Nanev Slavov; Vincent Louis Viala |
| EPI_ISL_1533704 | Santa Casa de Atibaia Pro Saude | Instituto Adolfo Lutz, Interdiciplinary Procedures Center, Strategic Laboratory | Caio Vinicius Dias Lopes; Claudia Regina Gonçalves; Claudio Tavares Sacchi; Erica Valessa Ramos Gomes; Karoline Rodrigues Campos; Leonardo Jose Tadeu de Araujo |
| EPI_ISL_2346041 | UBS JARDIM MARAGOGIPE | Instituto Butantan | Antonio Jorge Martins; Claudia Renata dos Santos Barros; David Schlesinger; Debora Botequio Moretti; Dimas Tadeu Covas; Elaine Cristina Marqueze; Elaine Vieira Santos; Evandra Strazza Rodrigues; Heidge Fukumasu; Jayme Augusto de Souza-Neto; José Salvatore Leister Patané; Luiz Alcantara; Luiz Lehmann Coutinho; Maria Carolina Elias; Maurício Lacerda Nogueira; Rafael dos Santos Bezerra; Raul Machado Neto; Rejane Maria Tommasini Grotto; Ricardo Haddad; Sandra Coccuzzo Sampaio Vessoni; Simone Kashima; Svetoslav Nanev Slavov; Vincent Louis Viala |
| EPI_ISL_2345985 | UBS JARDIM ODETE | Instituto Butantan | Antonio Jorge Martins; Claudia Renata dos Santos Barros; David Schlesinger; Debora Botequio Moretti; Dimas Tadeu Covas; Elaine Cristina Marqueze; Elaine Vieira Santos; Evandra Strazza Rodrigues; Heidge Fukumasu; Jayme Augusto de Souza-Neto; José Salvatore Leister Patané; Luiz Alcantara; Luiz Lehmann Coutinho; Maria Carolina Elias; Maurício Lacerda Nogueira; Rafael dos Santos Bezerra; Raul Machado Neto; Rejane Maria Tommasini Grotto; Ricardo Haddad; Sandra Coccuzzo Sampaio Vessoni; Simone Kashima; Svetoslav Nanev Slavov; Vincent Louis Viala |
| EPI_ISL_2345596 | UBS MARCIA CRISTIANE DA SILVA DE OURO VERDE | Instituto Butantan / ESALQ-Piracicaba | Antonio Jorge Martins; Claudia Renata dos Santos Barros; David Schlesinger; Debora Botequio Moretti; Dimas Tadeu Covas; Elaine Cristina Marqueze; Elaine Vieira Santos; Evandra Strazza Rodrigues; Heidge Fukumasu; Jayme Augusto de Souza-Neto; José Salvatore Leister Patané; Luiz Alcantara; Luiz Lehmann Coutinho; Maria Carolina Elias; Maurício Lacerda Nogueira; Rafael dos Santos Bezerra; Raul Machado Neto; Rejane Maria Tommasini Grotto; Ricardo Haddad; Sandra Coccuzzo Sampaio Vessoni; Simone Kashima; Svetoslav Nanev Slavov; Vincent Louis Viala |
| EPI_ISL_2758673, EPI_ISL_2758682 | UEL | IPEC Guarapuava | NAPI-Genômica (Novos Arranjo de Pesquisa e Inovação em Genômica): Ademar Dantas da Cunha Júnior Adriano Ferrasa Adriano Mondini Aldo Przybysz Alessandra Lourenço Cecchini Armani Alex Sandro Jorge Alexandra Ivo de Medeiros Alexandre Maller Aline Cristina Batista Rodrigues Johann Ana Lucia Ferreira Ana Marisa Fusco Almeida Anderson Joel Martino Andrade André Luís Laforga Vanzela Andrea Duarte Doetzer Andrea Name Colado Simao Andressa Pereira de Souza Anelisa Ramão Angelica Beate Winter Boldt Anna Hermínia Castro Gomes de Amorim Anna Silvia Penteado Setti da Rocha Antonio Camilo da Silva Filho Antonio Stabelini Neto Arthur Hirata Bertachi Barbara Mendes Paz Chao Betty Cristiane Kuhn Bruno Ambrozio Galindo Bruno Ribeiro Cruz Camilla Reginatto De Pierri Carla Fredrichsen Moya Araujo Carla Fredrichsen Moya Araujo Carlos Alberto Oliveira de Biagi Junior Carlos Augusto Nassar Carlos Eduardo Buss Carlos Gilberto Carlotti Junior Carlos Henrique Schneider Carolina Panis Carolina Weigert Galvão Caroline de Jesus Coelho Donha Caroline Guisantes de Salvo Toni Caryna Eurich Mazur Catiuscie Cabreira da Silva Tortorella Celso F. D. Doliveira Cesar Luiz Boguszewski Christiane Pienna Soares Chung Man Chin Claudia Moro Cleverson Busso Cristiane Cominetti Daiane Priscila Simão-Silva Dalila Luciola Zanette Daniel de Paula Daniel de Paula Daniel Rech Daniela Fiori Gradia Daniela Pretti da Cunha Tirapelli Daniela Viganó Zanoti Jeronymo Daniele Ukan Danielle Malheiros Ferreira Danielle Venturini Deborah Catharine de Assis Leite Deivid Calebe de Souza Dennis Armando Bertolini Edenir Inez Pamero Edna Maria Vissoci Reiche Edson Roberto Arpini Miguel Eduardo José de Almeida Araújo Eliana Carolina Vespero Eliandro Reis Tavares Elza Kimura Grimshaw Emanuel Maltempi de Souza Emanuele Cristina Gustani Buss Émerson Carraro Emiliana Cristina Melo ENILze Maria de Souza Fonseca Ribeiro Enilze Maria de Souza Fonseca Ribeiro Erika Izumi Erika Seki Kioshima Cotica Evani Marques Pereira Fabio Negretti Fábio Rodrigues Ferreira Seiva Felipe Dunin dos Santos Felipe Tuon Fernanda Andreia Rosa Fernanda Cestaro Prado Cortez Fernanda Ivanski Fernanda Maris Peria Flavia Regina Oliveira de Barros Franciele Aní Caovilla Follador Franciele Mara Lucca Zanardo Bohm Francinete Ramos Campos Fulviana Silva Nishiyama GABRIEL RIBEIRO CORDEIRO Gabriela Datsch Bennemann Gisele Santos de Oliveira Glaucio Valdameri Glauco Akelinghton Freire Vitiello Glauco Vieira Miranda Glaura Scantamburlo ALves Fernandes Guilherme Ferreira Silveira Gustavo Bianchini Porfírio Gustavo Lenci Marques Hélito Volpato |

| | | | |
|---|---|---|---|
| | | | Hildebrando Masshiro Nagai Huei Diana Lee Ilce Mara de Syllos Cólus Iris Rabinovich Israel Gomy Jackson Kawakami Jacques Duílio Brancher Jaime Luís Lopes Rocha Jaqueline Carvalho de Oliveira Jean Henrique da Silva Rodrigues Jean Leandro dos Santos Jeane Eliete Laguila Visentainer João Paulo Bianchi Ximenez Joaquim Manoel da Silva Jociani Ascari Joel Donazzolo Jorge Luis Maria Ruiz Jose Knopfholz José Luis da Conceição Silva José Sebastião dos Santos Joseane Carla Schabarum Juliana Cheleski Wiggers Juliana Mara Serpeloni Juliana Morini Küpper Cardoso Perseguini Karen Brajão de Oliveira Karin Braun Prado Karine Aparecida de Lima Katiany Rizzieri Caleffi Ferracioli Katiuscia de Oliveira Francisco Gabriel Kelvinson Fernandes Viana Larissa Beatriz Cossalter Larissa Danielle Bahls Pinto Laurival Antonio Vilas Boas Léia Carolina Lucio Líbero Mezzadri Neto Ligia Carla Faccin Galhardi Lirane Elize Defante Ferreto Luciana Furlaneto Maia Luciana Oliveira de Fariña Luciana Reis Azevedo Alanis Luciane Regina Cavalli Lucy Megumi Yamauchi Lioni Luis Paulo Gomes Mascarenhas Luis Paulo Gomes Mascarenhas Luís Paulo Mascarenhas Lupe Furtado Alle Lyvia Regina Biagi Silva Bertachi Mara Antonia Ramos Costa Mara L. Cordeiro Marcela Maria Birolim Marcelo Ricardo Vicari Marcia Edilaine Lopes Consolaro Marcia Holsbach Beltrame Marcia Regina Eches Perugini Marcos Abdo Arbex Marcos Pileggi MARCOS TADEU GRZELCZAK Marcus Peikriszwili Tartaruga Maria Angelica Ehara Watanabe Maria Antonia Ramos Costa Maria Claudia Gross Maria José Soares Mendes Giannini Maria Leandra Terencio Maria Lúcia Bonfleur Maria Luiza Guimarães de Oliveira Maria Luiza Petzl-Erler Mariana Abe Vicente Cavagnari Marina Kimiko Kadowaki Marise Fonseca dos Santos Marla Karine Amarante Maurício Turkiewicz Mauro Antonio Alves Castro Michel Rodrigo Zambrano Passarini Michele Potrich Michelle Orane Schemberger Milena Massumi Kozonoe Mônica Degraf Cavallin Monica Tereza Suldofski Mucio Luiz de Assis Cirino Nadia Graciele Krohn Najeh Maissar Khalil Nédia de Castilhos Ghisi Neide Tomimura Costa Neiva Leite Neyva Maria Lopes Romeiro Patrícia Amâncio da Rosa Patricia Dayane Carvalho Schaker Patricia Oehlmeyer Nassar Patrícia Savio de Araújo-Souza Patricia Silva Lucio Paulo Henrique Couto Souza Paulo Roberto Donadio Percy Nohama Quirino Alves de Lima Neto Rafael Deminice Rafael dos Santos Bezerra Raquel Alves dos Santos Renan Manozzo Galante Renata Ernlund Freitas de Macedo Rita de Cássia Garcia Simão Roberta Losi Guembarovski Roberto H. Herai Roberto Rosati Rodrigo Ferreira Rodrigo Rodrigues Matiello Rogério Neri Shinsato Rogério Pincela Mateus Rosane Aparecida Ribeiro Rosilene Fressatti Cardoso Rosilene Fressatti Cardoso Sandra Mara Guse Scós Venske Selene Elifio Esposito Sérgio Ossamu Ioshii Silvana Giuliatti Silvia Mara de Souza Halick Silvio Henrique Maia de Almeida Simone Neumann Wendt Spencer Luiz Marques Payão Stefan Wolanski Negrão Stephane Janaina de Moura Escobar Sueli Fumie Yamada Ogatta SUELI PERCIO QUINAIA Taciane Finatto Tatiana Mayumi Veiga Iriyoda Tayza Katelline Danilau Ostroski Tony Alexander Hild Valeria Valente Vanessa Nascimento Kozak Vanessa Santos Sotomaior Victor Breno Pedrosa Victoria Zeghbi Cochenski Borba Vivian Rotuno Moure Valdameri Wander Rogerio Pavanelli Weber Cláudio Francisco Nunes da Silva Willian Augusto de Melo Yohandra Reyes Torres |
| EPI_ISL_2494174 | UNIDADE MISTA DE SAUDE RAFARD | Instituto Butantan | Antonio Jorge Martins; Claudia Renata dos Santos Barros; David Schlesinger; Debora Botequio Moretti; Dimas Tadeu Covas; Elaine Cristina Marqueze; Elaine Vieira Santos; Evandra Strazza Rodrigues; Heidge Fukumasu; Jayme Augusto de Souza-Neto; José Salvatore Leister Patané; Luiz Alcantara; Luiz Lehmann Coutinho; Maria Carolina Elias; Maurício Lacerda Nogueira; Rafael dos Santos Bezerra; Raul Machado Neto; Rejane Maria Tommasini Grotto; Ricardo Haddad; Sandra Coccuzzo Sampaio Vessoni; Simone Kashima; Svetoslav Nanev Slavov; Vincent Louis Viala |
| EPI_ISL_2445277 | UPA VILA SANTA CATARINA | Instituto Butantan | Antonio Jorge Martins; Claudia Renata dos Santos Barros; David Schlesinger; Debora Botequio Moretti; Dimas Tadeu Covas; Elaine Cristina Marqueze; Elaine Vieira Santos; Evandra Strazza Rodrigues; Heidge Fukumasu; Jayme Augusto de Souza-Neto; José Salvatore Leister Patané; Luiz Alcantara; Luiz Lehmann Coutinho; Maria Carolina Elias; Maurício Lacerda Nogueira; Rafael dos Santos Bezerra; Raul Machado Neto; Rejane Maria Tommasini Grotto; Ricardo Haddad; Sandra Coccuzzo Sampaio Vessoni; Simone Kashima; Svetoslav Nanev Slavov; Vincent Louis Viala |
| EPI_ISL_2345984 | USF JARDIM PIRATININGA ITAQUAQUECETUBA | Instituto Butantan | Antonio Jorge Martins; Claudia Renata dos Santos Barros; David Schlesinger; Debora Botequio Moretti; Dimas Tadeu Covas; Elaine Cristina Marqueze; Elaine Vieira Santos; Evandra Strazza Rodrigues; Heidge Fukumasu; Jayme Augusto de Souza-Neto; José Salvatore Leister Patané; Luiz Alcantara; Luiz Lehmann Coutinho; Maria Carolina Elias; Maurício Lacerda Nogueira; Rafael dos Santos Bezerra; Raul Machado Neto; Rejane Maria Tommasini Grotto; Ricardo Haddad; Sandra Coccuzzo Sampaio Vessoni; Simone Kashima; Svetoslav Nanev Slavov; Vincent Louis Viala |
| EPI_ISL_1858977, EPI_ISL_1858979 | Unidade de apoio ao diagnóstico da COVID – UNADIG | Bioinformatics Laboratory / LNCC | Alessandra P Lamarca; Alexandra L Gerber; Amilcar Tanuri; Ana Paula de C Guimarães; Ana Tereza R Vasconcelos; Andréa Cony Cavalcanti; Caio Luiz Pereira Ribeiro; Cassia Alves; Claudia Maria Braga de Mello; Cristiane Gomes da Silva; Diana Mariani; Douglas Terra Machado; Flávio Dias da Silva; Leandro Magalhães de Souza; Liliane Cavalcante; Luiz G P de Almeida; Marcio Henrique de Oliveira Garcia; Mario Sergio Ribeiro; Ronaldo da Silva F Jr; Silvia Carvalho; Thais Felix Cruz |
| EPI_ISL_584625 | University College London Hospital | COVID-19 Genomics UK (COG-UK) Consortium | Catherine Houlihan; Dan Frampton; Judith Heaney; Matthew Byott; Moira Spyer and Eleni Nastouli; Stuart Kirk |
| EPI_ISL_2210295 | VIGILANCIA SANITARIA E VIG EPIDEMIOLOGICA DE ITAPEVI | Instituto Butantan | Antonio Jorge Martins; Claudia Renata dos Santos Barros; David Schlesinger; Debora Botequio Moretti; Dimas Tadeu Covas; Elaine Cristina Marqueze; Elaine Vieira Santos; Evandra Strazza Rodrigues; Heidge Fukumasu; Jayme Augusto de Souza-Neto; José Salvatore Leister Patané; Luiz Alcantara; Luiz Lehmann Coutinho; Maria Carolina Elias; Maurício Lacerda Nogueira; Rafael dos Santos Bezerra; Raul Machado Neto; Rejane Maria Tommasini Grotto; Ricardo Haddad; Sandra Coccuzzo Sampaio Vessoni; Simone Kashima; Svetoslav Nanev Slavov; Vincent Louis Viala. |
| EPI_ISL_2345885 | VIGILANCIA SANITARIA E VIG EPIDEMIOLOGICA DE ITAPEVI | Instituto Butantan / FZEA-USP-Pirassununga | Antonio Jorge Martins; Claudia Renata dos Santos Barros; David Schlesinger; Debora Botequio Moretti; Dimas Tadeu Covas; Elaine Cristina Marqueze; Elaine Vieira Santos; Evandra Strazza Rodrigues; Heidge Fukumasu; Jayme Augusto de Souza-Neto; José Salvatore Leister Patané; Luiz Alcantara; Luiz Lehmann Coutinho; Maria Carolina Elias; Maurício Lacerda Nogueira; Rafael dos Santos Bezerra; Raul Machado Neto; Rejane Maria Tommasini Grotto; Ricardo Haddad; Sandra Coccuzzo Sampaio Vessoni; Simone Kashima; Svetoslav Nanev Slavov; Vincent Louis Viala |
| EPI_ISL_430533 | Victorian Infectious Diseases Reference Laboratory (VIDRL) | Microbiological Diagnostic Unit Public Health Laboratory and Victorian Infectious Diseases Reference Laboratory, The Peter Doherty Institute for Infection and Immunity | Caly L.; Druce J.; Sait, M.; Schultz M.; Seemann T.; Sherry, N. |
| EPI_ISL_510771 | Viollier AG | Department of Biosystems Science and Engineering, ETH Zürich | Christian Beisel; Christiane Beckmann; Christoph Noppen; Elodie Burcklen; Ina Nissen; Ivan Topolsky; Maurice Redondo; Natascha Santacroce; Niko Beerenwinkel; Noemie Santamaria de Souza; Olivier Kobel; Pedro Ferreira; Philipp Jablonski; Sarah Nadeau; Sophie Seidel; Susana Posada-Céspedes; Tanja Stadler; Tobias Schär |
| EPI_ISL_402124 | Wuhan Jinyintan Hospital | Wuhan Institute of Virology, Chinese Academy of Sciences | Ding-Yu Zhang; Hao-Rui Si; Lei Zhang; Peng Zhou; Xing-Lou Yang; Yan Zhu; Zhengli Shi |
| EPI_ISL_458146, EPI_ISL_524787, EPI_ISL_524796, EPI_ISL_524797, EPI_ISL_524798, EPI_ISL_524799, EPI_ISL_848598, EPI_ISL_848601, EPI_ISL_848615, EPI_ISL_848617, EPI_ISL_848619, EPI_ISL_848622, EPI_ISL_848624, EPI_ISL_848625, EPI_ISL_848628 | | | |
| see above | Evandro Chagas Institute | Evandro Chagas Institute | A.M.; Barbagelata; E.C.; E.M.A.; Ferreira; G.M.R; H.R; J.A.; Junior; K.C.; L.C.; L.S.; M.C.; Martins; P.S.; Pinheiro; Resque; Santos; Silva; Sousa; Sousa Junior; Viana; W.D.C.; da Silva |
