## Supplemental File for "A case series of SARS-CoV-2 reinfections caused by the variant of concern Gamma in Brazil"

**PCR amplification for the whole SARS-CoV-2 genome.**

We have previously published a protocol to generate high-quality near to full length genomes positions 47 to 29,835 or 99.6% of the SARS-CoV-2 reference sequence EPI_ISL_402124 [1]. We observed that some sequences were better amplified (e.g., stronger amplification of the final amplicon observed in agarose gel electrophoresis) using a combination of two reverse primers at the 3' end (**Supplementary table 2**). Additionally, some thermocycling conditions were adjusted (**Supplementary table 3**). Therefore, using this alternative protocol some sequences have 29,593bp, positions 47 to 29,644 (98.96%) of the EPI_ISL_402124.

**Supplementary table 3.** Primers strategy used for the whole-genome sequencing of the SARS-CoV-2 based on the strategy described previously [1] .

| Amplicon | Primer Name | Sequence 5`-3 | pb |
|---|---|---|---|
| 1 | 24_F | GTAACAAACCAACCAACTTTCGA | 3962 |
| | 3986_R | TTGTAACTTCTTCAACACAAGCTTT | |
| 2 | 3841_F | AATGAAGAGTGAAAAGCAAGTTGAA | 4125 |
| | 7966_R | TTGACACATAAGCTGACTGTAGTAA | |
| 3 | 7836_F | TAGACAACCTGAGAGCTAATAACAC | 4153 |
| | 11989_R | AAAGGCTTCAGTAGTATCTTTAGCT | |
| 4 | 11857_F | TGTAAAGTGCACATCAGTAGTCTTA | 4203 |
| | 16060_R | TAGGATGTTTAGTAAGTGGGTAAGC | |
| 5 | 15911_F | ATGATTATGTGTACCTTCCTTACCC | 4123 |
| | 20034_R | ACGGGCATTTCTAAATAAGTCTACT | |
| 6 | 19865_F | ATACTGTGATCTGGGACTACAAAAG | 4279 |
| | 24144_R | AAAACAGTAAGGCCGTTAAACTTTT | |
| 7 | 23997_F | CAAGCAAGAGGTCATTTATTGAAGA | 2036 |
| | 26032_R | GCTGGTAATAGTCTGAAGTGAAGTA | |
| 8 | 25827_F | TTTTCTTTGCTGGCATACTAATTGT | 3120 |

| | | | |
|---|---|---|---|
| | W_HU_1_28946R | CAAGCAGCAGCAAAGCAAGA | |
| 9 | 28155_F | ATTAATTGCCAGGAACCTAAATTGG | 1515 / 1707 |
| | 29672_R | ATGTGAGATTAAAGTTAACTACATCT | |
| | 29861_R | CTAAGAAGCTATTAAAATCACATGGG | |

**Supplementary table 4.** Thermocycling conditions to amplify the SARS-CoV-2 genomes.

| Number of Cycles | Steps | Temperature | Time |
|---|---|---|---|
| 1 | Initial denaturation | 98ºC | 30 seconds |
| | Denaturation | 98ºC | 10 seconds |
| 35 | Hybridization | **60ºC or 55ºC*** | 10 seconds |
| | Extension | 72ºC | 2 minutes |
| 1 | Final extension | 72ºC | 5 minutes |
| 1 | Hold | 4ºC | ∞ |

*All the amplicons 1 to 8 are amplified under the hybridization temperature 60ºC except amplicon 9 which is amplified under the hybridization temperature 55ºC
